# Cancer Diagnosis, Polygenic Risk, and Longevity-Associated Variants

**DOI:** 10.1101/2020.09.18.20197475

**Authors:** Laura H. Goetz, Janith Don, Andrew J. Schork, David Duggan, Nathan D. Price, Daniel S. Evans, Steve Cummings, Thomas Perls, Paola Sebastiani, Nicholas J. Schork

## Abstract

**Background:** Polygenic risk scores (PRS) have been developed to predict individual cancer risk and their potential clinical utility is receiving a great deal of attention. However, the degree to which the predictive utility of individual cancer-specific PRS may be augmented or refined by the incorporation of other cancer PRS, non-cancer disease PRS, or the protective effects of health and longevity-associated variants, is largely unexplored.

**Methods:** We constructed PRS for different cancers from public domain data as well as genetic scores for longevity (‘Polygenic Longevity Scores’ or ‘PLS’) for individuals in the UK Biobank. We then explored the relationships of these multiple PRS and PLS among those with and without various cancers.

**Results:** We found statistically significant associations between some PLS and individual cancers, even after accounting for cancer-specific PRS. None of the PLS in their current form had an effect pronounced enough to motivate clinical cancer risk stratification based on its combined use with cancer PRS. A few variants at loci used in the PLS had known associations with Alzheimer’s disease and other diseases.

**Conclusion:** Underlying heterogeneity behind cancer susceptibility in the population at large is not captured by PRS derived from analytical models that only consider marginal associations of individual variants with cancer diagnoses. Our results have implications for the derivation and calculation of PRS and their use in clinical and biomedical research settings.

**Impact:** Extensions of analyses like ours could result in a more refined understanding of cancer biology and how to construct PRS for cancer.

## INTRODUCTION

While cancer treatments and outcomes have improved significantly over the past 40 years, overall cancer incidence continues to rise.(1) In 2019, an estimated 16.9 million cancer survivors lived in the United States, a seven-fold increase since 1971.(2) Cancers are primarily diseases of aging, with over 80% of cancers occurring in individuals over age 50.(1) Since this proportion of the US population is projected to increase steadily over the next several decades,(3) there is an urgent need to address this impending public health and medical crisis through the development of strategies to decrease rates of cancers in aging adults. Developing appropriate strategies will require identifying individuals at risk for cancer and providing them with specific risk mitigation interventions that go beyond current prevention and screening recommendations. In this light, cancer risk assessment can be greatly enhanced by using genetic profiling. Both monogenic and polygenic influences play an important role in susceptibility to and pathogenesis of cancers, however, cancers in the aged are less likely to be associated with monogenic germline variants(4-6) The degree to which polygenic variants contribute specifically to aging-related cancer risk is the subject of much current research.(7,8)

GWAS have identified thousands of common genetic variants associated with many different types of cancer.(9,10) Although each individual variant has a small effect on risk, especially relative to those contributing to monogenic forms of cancer, their cumulative effects can be pronounced. These *polygenic effects* can make individual contributions to risk equal to that of the individual rare variants contributing to monogenic forms of hereditary cancer (11) and also modify or compound the risk of these monogenic driver variants(12-14). These common variant, polygenic effects are often summarized at the individual level as a polygenic risk score (PRS) that aggregates effects across multiple variants into a single estimate of the total cancer susceptibility burden in a given genome. Cancer PRS are derived in different ways, but often use simple weighted multiple regression and related prediction modeling techniques.(15-17). **Supplementary Table 1** provides a list of recent cancer PRS studies.

**Table 1:**
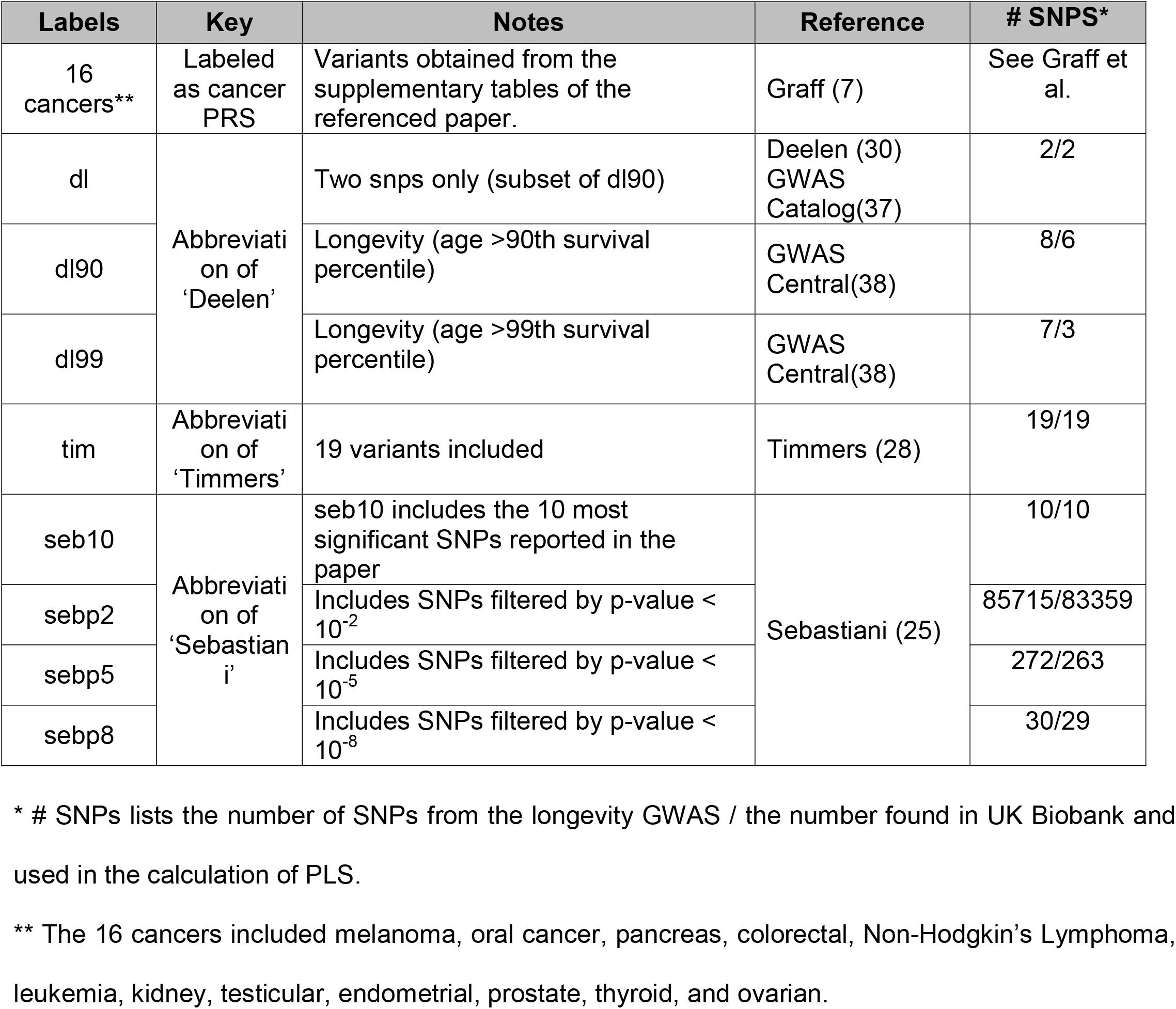
Key to PRS/PLS labels with references of summary statistics used for each score.

| Labels | Key | Notes | Reference | # SNPS* |
| --- | --- | --- | --- | --- |
| 16 cancers** | Labeled as cancer PRS | Variants obtained from the supplementary tables of the referenced paper. | Graff (7) | See Graff et al. |
| dl | Abbreviation of 'Deelen' | Two snps only (subset of dl90) | Deelen (30)<br>GWAS Catalog(37) | 2/2 |
| dl90 |  | Longevity (age >90th survival percentile) | GWAS Central(38) | 8/6 |
| dl99 |  | Longevity (age >99th survival percentile) | GWAS Central(38) | 7/3 |
| tim | Abbreviation of 'Timmers' | 19 variants included | Timmers (28) | 19/19 |
| seb10 | Abbreviation of 'Sebastiani' | seb10 includes the 10 most significant SNPs reported in the paper | Sebastiani (25) | 10/10 |
| sebp2 | | Includes SNPs filtered by p-value < $10^{-2}$ | | 85715/83359 |
| sebp5 | | Includes SNPs filtered by p-value < $10^{-5}$ | | 272/263 |
| sebp8 | | Includes SNPs filtered by p-value < $10^{-8}$ | | 30/29 |

\* # SNPs lists the number of SNPs from the longevity GWAS / the number found in UK Biobank and used in the calculation of PLS.
\*\* The 16 cancers included melanoma, oral cancer, pancreas, colorectal, Non-Hodgkin's Lymphoma, leukemia, kidney, testicular, endometrial, prostate, thyroid, and ovarian.

Interest in PRS grew from developing models that could discriminate specific cancer cases from controls, but has now expanded into multiple other areas. For example, PRS have been used to explore genetic, as opposed to phenotypic, correlations between cancers to better characterize shared genetic determinants of different cancers;(8,18) correlations between the PRS themselves for different cancer-related phenotypes;(19) the pleiotropic effects of variants across multiple different cancers;(20) and the causal effects of various measurable and potentially modifiable genetically-mediated factors on cancer via, e.g., Mendelian Randomization tests.(21-23) This work suggests that being susceptible to one type of cancer or phenotype can influence susceptibility - positively or negatively - to another type.(21,24) The degree to which this is the case is an open question.

Fundamental processes mediating cancer susceptibility that are shared across cancers are also likely to mediate susceptibility to other diseases and conditions beyond cancer. Such processes are quite likely to be associated with aging and longevity, given that the manifestation of many common chronic diseases is age-related. By bringing together genetic insights into multiple cancers, diseases, aging and longevity, one might be able to develop not only better prediction models through, e.g., risk stratification, but also more insight into the genetic etiology of cancer. Identifying variants associated with health span and longevity is challenging. Different definitions of longevity are used, sample sizes in studies focusing on extreme longevity are small, and the actual heritability of human lifespan is debated, as is the need to account for environmental factors in relevant studies.(25-27) Large-scale analyses of long-lived cohorts have enabled pooled analysis of multiple GWAS, as has been the focus of the National Institute on Aging’s Longevity Consortium (LC), the Integrated Longevity Omics (ILO), and Long Life Family Study (LLFS) projects.(25) The results of these studies have yet to be integrated with studies of various diseases. GWAS focused on genetic ‘surrogates’ for individual lifespan, such as parental lifespan, have also been pursued.(28,29) Although not all studies are consistent, many of these analyses suggest that long lived individuals possess different disease PRS profiles from short lived individuals.(30-33) Some individuals possessing a genetic background consistent with a long-life – i.e., having an elevated Polygenic Longevity Score (PLS) – may have inherited genetic variants that protect them from cancer and other diseases. However, these variants may not have individual protective effects on cancer pronounced enough to be detected as significantly associated with a cancer diagnosis in cancer-specific GWAS.(34)

We explored the relationships among PRS built from published GWAS results and PLS derived from different longevity-focused GWAS and their relationships to specific cancer diagnoses, general cancer risk, and age of cancer onset in the UK Biobank.(35,36) We also considered the development and performance of modified cancer risk prediction models that included longevity associated variants. We find that although many PLS are associated with cancers at some level, these effects are not pronounced enough to radically impact the use of cancer PRS prediction models. We do, however, argue that the relationship between longevity, cancers and disease protection needs greater exploration, especially given that some of the variants used in the cancer-associated PLS are strongly negatively associated with other diseases, like Alzheimer’s disease, raising important biological questions.

## METHODS

### Data Sources

We calculated polygenic risk scores (PRS) for 16 cancers using publicly available data,(7,37,38) as well as polygenic longevity scores (PLS) using 8 different longevity measures derived from different publicly available longevity GWAS summary statistics and different p-value thresholds (**Table 1**; SNPs and weights used for each PLS are provided in the supplementary material).

PLS were labeled according to the source reference for the summary statistics (**‘dl’, ‘dl90’, ‘tim’**, etc. See Table 1). We note that some PLS were based on a subset of SNP markers identified from the various longevity-focused GWAS because there were no reported effect sizes and some were not among those in version-3 of the UK Biobank total SNP list, including imputed SNPs. All PRS and PLS were computed on each of the individuals in the UK Biobank with genotype calls from the UK Biobank version-3 genotype data using the linear scoring function in PLINK v2 software and published PRS effect sizes.(36,39,40) We restricted our analyses to White British individuals (based on UK Biobank Data-Field 21000) as the PRS and PLS we used were derived from individuals with Northern European ancestry and the simultaneous study of multiple ancestries can confound PRS inference.

### Logistic Regression Modeling

We used logistic regression to fit a number of prediction models to the UK Biobank data, with a specific cancer diagnosis or cancer PRS as a dependent variable and age, sex, PLS and the PRS for other cancer diagnoses as independent variables. For these analyses we included the first 10 principal components (PC) from the genome wide genetic relationship matrix (GRM) of the participants, although there is debate about the need or meaningfulness of including PCs in relevant analyses when the focus is on polygenic effects.(41-44) We also carried out stepwise logistic regression, using both forward and backward steps in a ‘both-way’ approach to calculate the best model, keeping independent variables in models in each step based on the Akaike information criterion (AIC) until no improvements in the model occurred. More details about the various models are described in corresponding sections in the Results section. All analyses were carried out using R version 3.6.3.

### Survival Analyses

We performed survival analyses on each of the 16 cancers and for ‘any-cancer’ diagnosis using age-of-onset information as the dependent variable and an individual’s age at assessment, if that individual was not diagnosed with cancer, as a censoring time variable. The UK Biobank recruited individuals between the ages of 40 and 70. We therefore did not include any cancer age-of-onsets that were reported after age 70 in follow-up studies with UK Biobank participants. Survival analyses were pursued using both the Kaplan-Meier models coupled with log-rank tests of differences in survival curves as a function of PLS, and Cox proportional hazards models with the first 10 PCs from the GRM (see above), cancer PRS and PLS as independent variables as implemented in the ‘survival’ R package.(45)

### Prediction Evaluation

We evaluated Receiver-Operator Characteristic (ROC) curves for each logistic regression model fit with cancer diagnosis as a dependent variable. We computed the Area-Under-the-Curve statistic (AUC) from the ROC analyses to gauge the performance of our logistic regression-based prediction models with age, sex, cancer PRS and PLS as independent variables. The R package pROC(46) was used for the relevant analyses.

## RESULTS

### Evidence for a shared genetic basis for different cancers

We first considered evidence that there might be a shared genetic basis to susceptibility to all cancers. We note that even if there is no evidence for a shared genetic basis for susceptibility across all or even most cancers, this does not preclude the possibility that one or a few cancers may share genetically-mediated determinants with longevity. Since individuals who live a long life did not die from cancer at earlier ages by definition, this suggests some degree of overlap in genetic determinants between cancers – and all diseases – and longevity (i.e., it may be that individuals who live a long life likely do not have genetic variants contributing to cancer susceptibility to the same degree as individuals who die earlier from cancer). We explored evidence for common genetic factors among cancers in three ways: 1. we considered studies in the literature whose results were consistent with a shared genetic basis for cancers; we analyzed data in the UK Biobank to test the hypothesis of a shared genetic basis; and 3. we explored the consistency between genetic correlations and PRS correlations among cancers.

**Supplementary Table 2** shows the number of individuals in the UK Biobank diagnosed with one cancer and individuals with at least two different cancers. There were only 45 individuals with 3 or more cancers and there was a maximum of 4 cancers observed for any individual. **Supplementary Table 3** depicts the AUC statistics from logistic regression models with specific cancer diagnosis as a dependent variable and age as well as other cancer diagnoses as independent variables, chosen based on stepwise logistic regression (see Methods). We note that some cancers are less frequent than others and as such some of these logistic regression analyses were likely underpowered, despite the large overall sample size of the UK Biobank. Both **Supplementary Tables 2 and 3** suggest that many cancer diagnoses are significantly associated with, and predictive of, other cancer diagnoses. Many of the relationships between cancers that we identified are well known: e.g., breast and ovarian cancer; endometrial and colorectal cancer; but some are not: e.g., bladder and breast cancer. We acknowledge and emphasize that the associations between cancers could be, at least in part, attributable to environmental factors that contribute to both cancers.

**Table 2.** AUC values derived for ROC analyses of the results of logistic regression modeling with specific cancer diagnosis as a dependent variable and the corresponding PRS for that cancer, PRS of the other cancers, age, and the different PLS as independent variables, including the first 10 Principal Components(PC) from the whole genome genetic relationship matrix (see Methods). We fit three models to determine the significance of the contribution of PLS: a ‘trivial model’ with just age as an independent variable; a ‘basic’ model with age and the cancer-specific PRS as independent variables; and a ‘final’ model in which age, the cancer-specific PRS, the PRS of the other cancers, and the eight PLS were independent variables subjected to forward and backward stepwise logistic regression with age fixed in all models. P value column corresponds to the most significant p values for the ‘final’ AUC model. The significant ‘final’ AUC values are shown in bold.

| Cancer | Number of Cases | AUC (trivial) | AUC (basic) | AUC (final) | p value | Additional PRS/PLS in the Final Model |
| --- | --- | --- | --- | --- | --- | --- |
| **MALES** |  |  |  |  |  |  |
| Bladder | 835 | 0.7046 | 0.7202 | 0.7253 | ns | lung, dl99, kidney, thyroid |
| Colon | 835 | 0.7047 | 0.7340 | **0.7369** | 0.006 | dl90, oral, prostate |
| Thyroid | 90 | 0.5366 | 0.6598 | 0.6742 | ns | kidney, tim |
| Melanoma | 1492 | 0.6031 | 0.6312 | 0.6312 |  | - |
| Kidney | 377 | 0.6366 | 0.6488 | 0.6500 | ns | sebp2 |
| Leukemia | 397 | 0.6199 | 0.6652 | 0.6710 | ns | bladder, colon, melanoma |
| Lung | 227 | 0.6886 | 0.6889 | 0.6961 | ns | prostate, colon, kidney |
| NHL | 513 | 0.5875 | 0.6134 | 0.6165 | ns | kidney |
| Oral | 293 | 0.6312 | 0.6336 | 0.6476 | ns | dl90, lung, melanoma, tim |
| Pancreas | 49 | 0.6900 | 0.6903 | **0.7416** | 0.0027 | lung, dl99, tim, thyroid |
| Prostate | 3700 | 0.7480 | 0.8032 | **0.8036** | 0.05 | kidney, melanoma, sebp8+ testicular, pancreas, NHL |
| Testicular | 774 | 0.5803 | 0.7284 | 0.7296 | ns | dl99, bladder |
| **FEMALES** |  |  |  |  |  |  |
| Bladder | 302 | 0.7019 | 0.7379 | **0.7462** | 0.003 | colon, thyroid, dl99, tim, NHL, leukemia |
| Colon | 638 | 0.6937 | 0.7277 | **0.7293** | 0.036 | dl, pancreas |
| Thyroid | 330 | 0.5325 | 0.6356 | 0.6406 | ns | endometrium, dl99 |
| Melanoma | 2101 | 0.5730 | 0.6189 | 0.6201 | ns | kidney, endometrium, oral |
| Kidney | 219 | 0.6360 | 0.6404 | 0.6603 | ns | NHL, lung, endometrium, melanoma, pancreas |
| Leukemia | 276 | 0.5955 | 0.6306 | 0.6377 | ns | thyroid, p2, cervix |
| Lung | 174 | 0.6854 | 0.6906 | 0.7009 | ns | colon, ovarian, kidney |
| NHL | 383 | 0.5897 | 0.6229 | 0.6329 | ns | Leukemia, bladder, lung |
| Oral | 184 | 0.6039 | 0.6050 | **0.6307** | 0.0038 | sebp8, sebp2, cervix, sebp5 |
| Pancreas | 34 | 0.6044 | 0.6138 | 0.6559 | ns | thyroid, p2 |
| Cervix | 3569 | 0.5483 | 0.5674 | **0.5695** | 0.04 | lung, p2, oral |
| Ovarian | 755 | 0.6019 | 0.6099 | 0.6130 | ns | thyroid, endometrium, breast |
| Breast | 10091 | 0.6232 | 0.6724 | **0.6732** | 0.0143 | colon, oral, p5, dl99, tim, endometrium, melanoma |
| Endometrium | 1137 | 0.6693 | 0.6741 | **0.6774** | 0.0047 | Leukemia, kidney, dl90, dl99, cervix |

**Table 3.** AUC values derived from ROC curve analysis of logistic regression models in which all 16 cancer diagnoses were combined and treated as a dependent variable and all cancer PRS, age, sex, and the PLS were considered independent variables. Independent variables for each case are shown inside the brackets. (Analysis results involving each individual cancer are provided in **Supplementary Table 7**.)

| Population | AUC (age) | AUC (age, cancer PRS) | AUC (age, cancer and PLS) |
| --- | --- | --- | --- |
| male | 0.66610 | 0.68355 | 0.68360 |
| female | 0.59181 | 0.61491 | 0.61518 |
| general | 0.64256 | 0.65164 | 0.65174 |

We explored the consistency between the strength of the genetic correlations and the strength of the PRS correlations, knowing that the different PRS used different sets of SNPs (see Discussion section). **Figure 1** provides a scatter plot of the estimates of genetic correlation obtained from Lindstrom et al(47) and corresponding PRS correlations computed on the UK Biobank subjects using the PRS of several cancers generated from the publicly available summary statistics (see Methods).(7)

**Figure 1:**
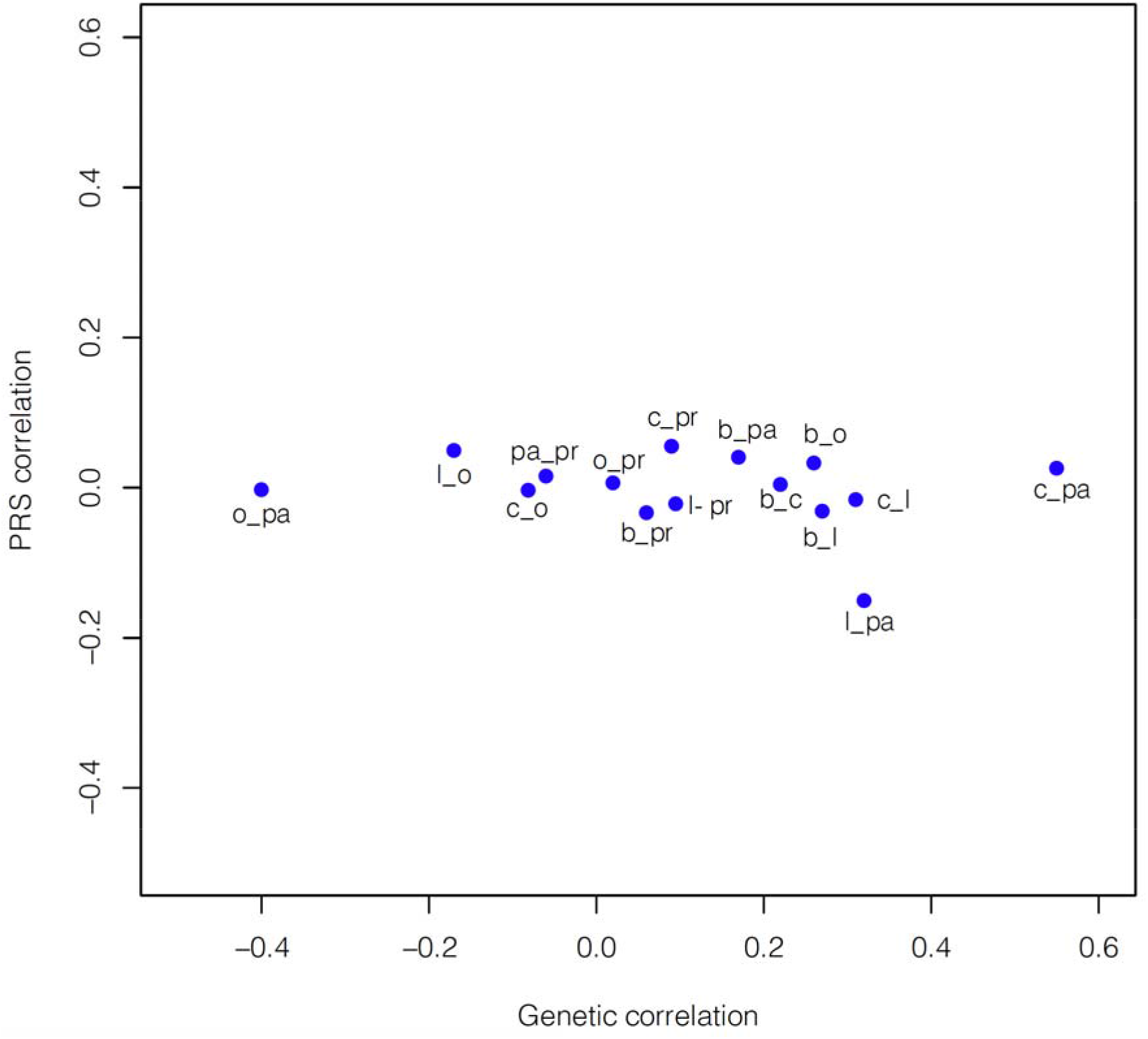
Scatter plot of the correlation strengths between 15 pairs of cancer PRS against the genetic correlations of those cancers based on the results of Lindstrom et al,(47) reflecting greater variation amongst genetic correlations and minimal variation amongst PRS correlations. (b=breast; c=colorectal; l=lung; o=ovarian; pa=pancreatic; pr=prostate)

We note that genetic and PRS correlations can be computed under different assumptions, using different methods with different overall orientations. For example, genetic correlations consider all the variants in a GWAS, whereas PRS correlations only use variants in the individual PRS. **Figure 1** suggests there is more variation in the genetic correlations than there is in the PRS correlations, perhaps reflecting the fact that the PRS correlations involve a small number of SNPs, even though many PRS correlations are significant. **Figure 2** provides a heatmap of the cancer PRS correlations for the UK Biobank participants and suggests that many cancers exhibit positive and negative PRS correlations.

**Figure 2:**
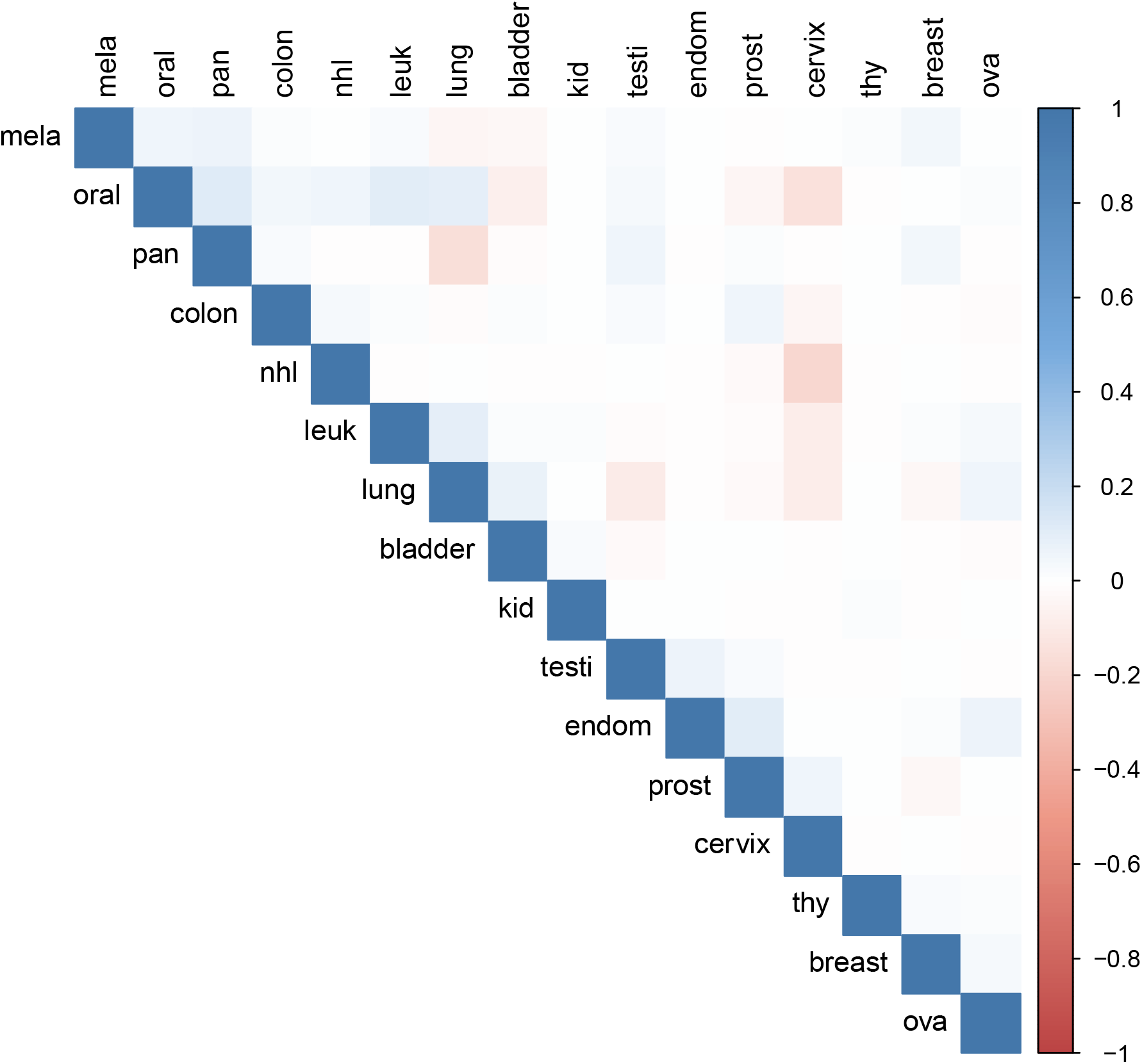
Colored correlation table reflecting the strength of the PRS correlations among 16 cancers. Red indicates a negative correlation and blue indicates a positive correlation. The stronger the correlation, the darker the color. The strongest correlations seen are a negative correlation between cervical cancer and Non-Hodgkin’s Lymphoma, cervical cancer and oral cancer, and lung cancer and pancreas cancer. (mela=melanoma, oral=oral cancer, pan=pancreas, colon=colorectal, nhl=Non-Hodgkin’s Lymphoma, leuk=leukemia, kid=kidney, testi=testicular, endom=endometrial, prost=prostate, thy=thyroid, ova=ovarian)

Note we included PRS correlations for cancers that are sex-specific for both sexes since such correlations may be meaningful biologically in terms of the pathways and underlying processes implicated. Since an obvious potential source for PRS correlations is overlap in the variants considered in the PRS being correlated, we tallied the number of overlapping variants used in the PRS we considered. The Supplementary Material provides a matrix with the number of SNPs in the summary statistics used to generate cancer PRS and number of intersecting SNPs across the cancers. More in-depth studies of the linkage-disequilibrium (LD) relationships between variants used in different PRS is called for to rule out more subtle sets of overlapping variant effects in different PRS.

### Associations Between Polygenic Longevity Score and Cancer

We assessed the relationships between the 8 different PLS listed in **Table 1** and cancer diagnosis, as well as cancer PRS. We note that one of our PLS, the PLS derived from the Timmers et al study, ‘tim,’(28) was derived from the UK Biobank data and studies involving actual UK Biobank participants, thus it should be kept in mind throughout our discussions that the study of this specific PLS could be confounded by an overfitting effect in our analyses. We took each cancer diagnosis as a dependent variable, and then considered the corresponding PRS for that cancer, PRS for the other cancers, age, and the different PLS as independent variables in stepwise logistic regression models. These analyses were pursued separately for each sex. The AUC statistics derived from these analyses consider: 1. age only; 2. age and cancer PRS; and 3. age, cancer PRS, and the PLS, and are summarized in **Table 2 and Supplementary Table 4**. (Specific models for each cancer with regression coefficients, etc. are available from the authors).

These tables suggest that PLS and other cancer PRS are associated with some cancers – Bladder, Colon, Pancreas, Prostate and Testicular cancer for men and Bladder and Colon for women – independently of the individual PRS for those cancers. However, the increase in the AUC score for these models when PLS are included are not large, and certainly not indicative of their potential for risk stratifying individuals in their current instantiation. We also computed correlations between the 16 cancer PRS and the 8 PLS. (**Supplementary Table 5**) We found small correlations, mostly negative as expected, although many were highly statistically significant. **Supplementary Table 6** provides a matrix of the correlations of each of the cancers with each of the eight PLS. Excluding the correlations for the ‘tim’ PLS, the largest negative correlation was between the ‘seb10’ PLS and the PRS for lung cancer and the largest positive correlation was between ‘sebp5’ and oral cancer.

### Age of Cancer Onset and Polygenic Longevity Score

Since the age-of-onset of cancers can be influenced by a number of factors, including genetic factors, we explored the association of the 8 PLS to cancer age-of-onset. We took information about the age-of-onset of each cancer in the UK Biobank and considered it in Cox proportional hazards models as well as a standard Kaplan-Meier (KM) survival analysis (see Methods). The age of individuals at the time they participated in the UK Biobank studies was used as a censoring age for individuals without a cancer diagnosis. **Supplementary Tables 7 and 8** provides the results of the Cox and KM analyses for each cancer and PLS, as well as analyses considering the age-of-onset of any cancer, and suggest that many cancers have an age-of-onset that is highly statistically significant in its association with one or another PLS, particularly the ‘**dl’**, ‘**dl90’** and ‘**dl99’** PLS. To depict the relationships between PLS and age-of-onset of cancers, we generated KM-based survival curves for individuals whose PLS were in different PLS distribution percentiles. **Figure 3** depicts the KM curves for analyses using age-of-onsets of any cancer diagnosis as well as for female breast cancer using the **‘dl’** PLS, contrasting individuals with **‘dl’** PLS in the upper and lower 10^th^ percentiles of the PLS scores among the UK Biobank participants. (See **Supplementary Figure 1** for KM curves for cervical cancer and PLS).

**Figure 3:**
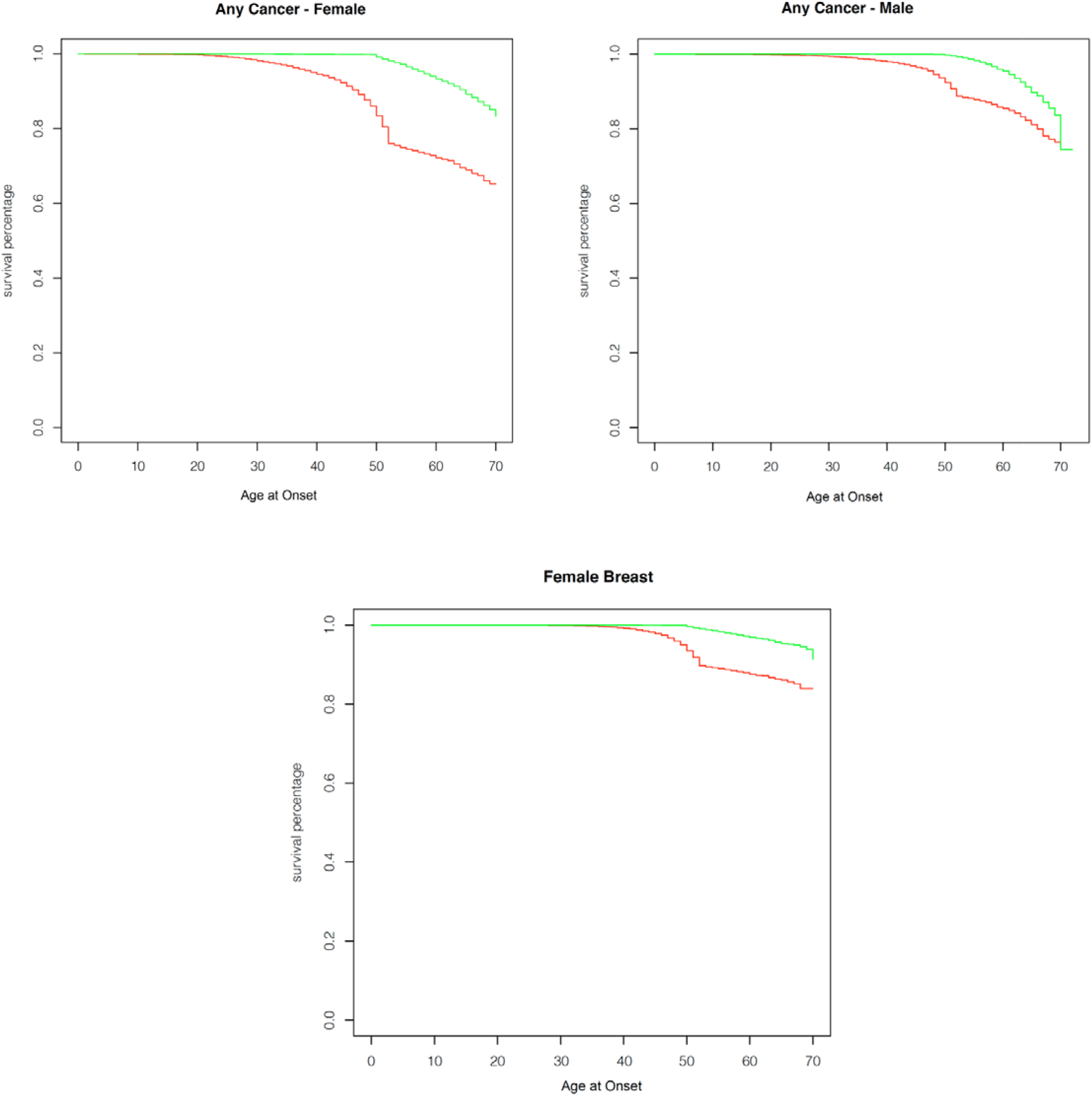
Kaplan-Meier curves showing effect of the **‘dl’** PLS on age-of-onset of select cancers. Red indicates individuals in the lower 10^th^ percentile of PLS and green indicates individuals in th 10^th^ percentile of PLS scores. Upper left panel: females with any cancer; Upper right panel upper males with any cancer. Lower panel: Females with breast cancer. All p-values for tests of the relationship of the percentile categories and cancer age-of-onset are p<10^−16^ and thus highly significant even after multiple comparisons corrections (see Methods).

### Cancer Prediction Modeling Performance

To determine the clinical utility of cancer prediction modeling that includes PLS in addition to cancer PRS (and age and sex), we computed Receiver Operator Characteristic (ROC) curves and the area-under-the-curve (AUC) statistic from logistic regressions with all the 16 cancer diagnoses combined and treated as a dependent variable (i.e., a dummy dependent variable was constructed whereby 1 = any-cancer diagnosis, and 0 = no cancer diagnosis) and all cancer PRS, age and sex as independent variables. We then added PLS as independent variables to the models and tested to see if the AUC was improved significantly, but did not find any statistically significant improvement (**Table 3**). We also carried out a similar analysis for each of the cancers; first using only age as an independent variable and then adding a single PLS. We also did not find significant improvements in AUCs (**Supplementary Table 9**). Thus, although PLS are significantly associated with cancer diagnoses and age at onset, their inclusion in clinical prediction models is not justified at this time.

## DISCUSSION

Many cancers share underlying etiological processes and factors (e.g., faulty DNA repair capacity, immune surveillance dysfunctions, etc.),(48) suggesting that the genetic determinants of these processes and factors mediate all or different subsets of cancers. The degree of overlap in the genetic determinants of cancers has been explored through the examination of the genetic correlations derived from individual GWAS results and data, as well as studies of the pleiotropic effects of individual genetic variants on multiple cancers.(7,18,20,49,50) In addition, many cancers have also been shown to be genetically correlated with non-cancer-related diseases.(24,51) These genetic correlations, possibly exacerbated by relevant environmental factors, are also quite likely to lead to cancer patients developing a second primary cancer (as opposed to recurrent cancers) or have comorbid illness. In this light, many other less obvious diseases have been found to be genetically correlated to specific cancers, such as psychiatric illness and lung and breast cancer.(52) Also, and perhaps more germane to our study of PRS and PLS, a recent study by Witte et al.(7) found that many variants associated with one form of cancer were also associated with another (e.g., exhibited pleiotropic effects across different cancer types) and that variants used in the PRS of one cancer are often included in the PRS of other cancers. They also found that PRS values were correlated for many pairs of cancers, as have many others.(8,18-20,53)

Since cancers can compromise lifespan, it is possible that individuals who live a long time: 1. do not have the variants that predispose to cancer to the same degree as individuals who have not lived a long time; 2. possess variants that protect them from the deleterious effects of cancer predisposing variants; 3. have been exposed to environmental factors (e.g., diet, lifestyle, etc.) that protect them from cancer in some way; or 4. had or have a combination of the above. Since many published studies provide evidence that genetic variants exist that predispose to a long life,(30,31,34) a good question concerns the degree to which such variants are absent from cancer PRS, or whether the possession of longevity-enhancing variants protects individuals against the deleterious effects of having an elevated cancer PRS. This is of particular interest given that cancer is primarily a disease of aging, and thus cancer’s genetically-mediated relationship to longevity may be pronounced. In addition, exploring and characterizing the positive genetic correlations between cancers, between cancers and other diseases, and the likely negative genetic correlations between cancers and longevity, can shed light on the pathogenesis of aging-related diseases, and also potential risk stratification strategies for identifying individuals at risk for a specific cancer, multiple cancers, disease sequelae and correlated morbidities, and complex disease trajectories whose interventions may be complicated.

Our study of the relationship between PLS and cancer diagnosis, age-of-onset and PRS in the UK Biobank suggests that although PLS are primarily negatively associated with many cancers, these associations are not strong enough to be used to modify risk assessments in clinical contexts in their current state. Our study used the UK Biobank data to evaluate associations between PLS, cancer PRS and cancer diagnoses and age-of-onsets, but considered PLS and cancer PRS for these analyses that were derived independently of the UK Biobank, with the exception of the ‘**tim’** PLS, to avoid overfitting. Despite this, there are several limitations to our study that should motivate further research. First, we did not consider an analysis involving the genetic basis of many modifiable risk factors that could impact susceptibility to, or protection from, cancer, such as diet, stress, exercise, smoking, etc. that themselves might be under genetic control.(54,55) Second, our study, like most GWAS-based studies, is only relevant to individuals of Northern European descent, and it is known that many cancers are more prevalent in individuals of non-European descent.(1) This is a well-documented criticism of PRS studies(56) and highlights the importance of conducting more diverse large-scale genomics research, inclusive of racially and ethnically diverse populations, such as the All-of-Us project, (57) before PRS (and PLS) can be exploited in a clinical setting.

Third, our analyses focused on common variants of the type used in most PRS calculations based on GWAS data and as such ignores rare variants that have been shown to enhance longevity and protect against diseases.(58-60) Fourth, most PRS have been derived from cross-sectional data (i.e., cases and controls) and yet are thought to be useful for both classifying individuals with and without cancer diagnoses, and predicting disease. Future analyses should consider the association of PLS and cancer development with longitudinal data that considers age, aging, long term exposure to environmental factors, and overall health trajectory to get a clearer picture of their interplay. Fifth, for reasons of scope, we did not consider detailed functional assessments of the individual genes harboring the variants, nor the linkage disequilibrium patterns they exhibit, in the various longevity and cancer PRS, which, if pursued in future studies, could shed light on the more basic biology behind the relationship between the longevity and cancer. We do, however, point out that many of the variants used to construct the 8 PLS we studied have been shown to be negatively correlated with other diseases, suggesting there are important biological relationships between PLS and PRS variants. For example, the ‘**dl’** PLS we studied is made up of only two SNPs, but those SNPs are known to be associated with Alzheimer’s disease (AD), suggesting a relationship between multilocus influences on AD and cancers.(61,62)[See the supplementary material for SNP information.)

An additional issue concerns why variants associated with longevity used in the PLS we studied are not included in cancer PRS if, in fact, the PLS are associated with cancer diagnosis and cancer PRS. Although there are a number of explanations for this which should motivate further study, we believe this is ultimately due to small marginal effects each variant used to construct a cancer PRS (or PLS) has on cancer susceptibility and the fact that the nearly the same score for a PRS can be obtained with different combinations of genotypes. Small individual effects make it unlikely that all biologically relevant variants are identified in studies used to derive PRS (or PLS) from a single finite data set. Also, there is likely great locus heterogeneity underlying cancer susceptibility and most diseases, as well as lifespan. Finally, cancer PRS are often trained on cases that possess a number of susceptibility variants, against controls that do not, and, as such, the variants used in PRS that are predictive of cancer diagnosis (or longevity) act as surrogates for the presence of others. Despite these issues, our results suggest that genetically-mediated factors that contribute to cancer susceptibility are indeed complex, but can be teased apart to some degree by leveraging insights into the genetic basis of not only different cancers, but also health-positive traits such as longevity. More detailed and focused studies of specific genetically-mediated factors that might contribute to both cancer risk reduction and longevity, including studies involving longitudinal data, are needed.

## Data Availability

All data is publicly available from the UK Biobank and/or the authors.

## ACKNOWLEDGEMENTS

This research was conducted with the UK Biobank Resource. The Project ID is 43036. Aspects of this work were funded in part by NIH grants UH2 AG064706, U19 AG023122, U24 AG051129, U24 AG051129-04S1; NSF grant (FAIN number) 2031819; Dell, Inc.; and the Ivy and Ottesen Foundations.

## FINANCIAL DISCLOSURES

The authors declare no financial conflicts

## Supplementary Material

**Supplementary Table 1.** Recent papers on polygenic risk scores and cancer∗

| Study | Year | Cancer Type | Number of SNPs | p value cutoff | Cases | Controls | AUC |
| --- | --- | --- | --- | --- | --- | --- | --- |
| Shieh(63)** | 2020 | Breast | 180 | $p < 5 \times 10^{-8}$ | 4658 | 7622 | 0.63 |
| Mavaddat(64) | 2019 | Breast | 313 | $p < 10^{-5}$ | 94,075 | 75,017 | 0.63 |
| Seibert(65) | 2018 | Prostate | 54 | $p < 10^{-7}$ | 18,868 | 12,879 | n/a |
| Yang(66) | 2018 | Ovarian | 96 | $p < 10^{-5}$ | 750 | 1428 | 0.60 |
| Loveday(67) | 2018 | Testicular | 37 | $p < 10^{-8}$ | 4167 | 12,368 | n/a |
| Lecarpentier(68) | 2017 | Male Breast | 88 | $p < 10^{-5}$ | 308 | 1469 | 0.59 |
| Lecarpentier(68) | 2017 | Prostate | 103 | $p < 10^{-5}$ | 255 | 1469 | 0.62 |
| Kuchenbaecker(69) | 2017 | Breast | 88 | $p < 10^{-8}$ | 12,127 | 11,336 | 0.56-0.58 |
| Kuchenbaecker(69) | 2017 | Ovarian | 17 | $p < 10^{-8}$ | 3093 | 20,370 | 0.58-0.63 |

\*Ancestry represented in the studies is predominantly European unless indicated.
\*\*Latinas and Latina-American Women

**Supplementary Tables 2a-b.** Matrices with cancer diagnosis by sex. Central diagonal contains the number of individuals diagnosed with an individual cancer. Corresponding non-diagonal positions contain the number of individuals diagnosed with a combination of cancers.

**2a) Male individuals with cancers**
|  | **blad** | **colon** | **thy** | **mel** | **pros** | **leuk** | **kid** | **pan** | **testi** | **oral** | **nhl** | **lung** |
| --- | --- | --- | --- | --- | --- | --- | --- | --- | --- | --- | --- | --- |
| *blad* | 835 | 6 | 1 | 9 | 58 | 1 | 9 | 0 | 8 | 3 | 3 | 4 |
| *colon* | 6 | 835 | 0 | 7 | 22 | 0 | 2 | 2 | 2 | 2 | 3 | 21 |
| *thy* | 1 | 0 | 90 | 2 | 2 | 0 | 1 | 0 | 1 | 0 | 1 | 0 |
| *mel* | 9 | 7 | 2 | 1492 | 39 | 5 | 6 | 1 | 11 | 5 | 9 | 3 |
| *pros* | 58 | 22 | 2 | 39 | 3700 | 11 | 22 | 0 | 9 | 5 | 11 | 11 |
| *leuk* | 1 | 0 | 0 | 5 | 11 | 397 | 2 | 0 | 2 | 0 | 5 | 1 |
| *kid* | 9 | 2 | 1 | 6 | 22 | 2 | 377 | 1 | 5 | 0 | 1 | 12 |
| *pan* | 0 | 2 | 0 | 1 | 0 | 0 | 1 | 49 | 0 | 0 | 0 | 0 |
| *testi* | 8 | 2 | 1 | 11 | 9 | 2 | 5 | 0 | 774 | 1 | 6 | 8 |
| *oral* | 3 | 2 | 0 | 5 | 5 | 0 | 0 | 0 | 1 | 293 | 2 | 2 |
| *nhl* | 3 | 3 | 1 | 9 | 11 | 5 | 1 | 0 | 6 | 2 | 513 | 1 |
| *lung* | 4 | 21 | 0 | 3 | 11 | 1 | 12 | 0 | 8 | 2 | 1 | 227 |

2b) Female individuals with cancers
|  | **blad** | **colon** | **brst** | **ova** | **thy** | **cer** | **mel** | **endom** | **leuk** | **kid** | **pan** | **oral** | **nhl** | **lung** |
| --- | --- | --- | --- | --- | --- | --- | --- | --- | --- | --- | --- | --- | --- | --- |
| *blad* | 302 | 4 | 21 | 2 | 1 | 8 | 1 | 4 | 0 | 5 | 0 | 0 | 1 | 2 |
| *colon* | 4 | 638 | 46 | 10 | 2 | 14 | 8 | 13 | 1 | 3 | 1 | 1 | 1 | 7 |
| *brst* | 21 | 46 | 10091 | 55 | 20 | 116 | 94 | 80 | 12 | 10 | 1 | 8 | 12 | 26 |
| *ova* | 2 | 10 | 55 | 755 | 0 | 21 | 8 | 28 | 0 | 1 | 0 | 0 | 2 | 8 |
| *thy* | 1 | 2 | 20 | 0 | 330 | 7 | 5 | 1 | 0 | 0 | 0 | 2 | 0 | 1 |
| *cer* | 8 | 14 | 116 | 21 | 7 | 3569 | 35 | 41 | 6 | 4 | 0 | 3 | 5 | 5 |
| *mel* | 1 | 8 | 94 | 8 | 5 | 35 | 2101 | 16 | 4 | 6 | 2 | 3 | 6 | 3 |
| *endom* | 4 | 13 | 80 | 28 | 1 | 41 | 16 | 1137 | 3 | 2 | 0 | 1 | 1 | 4 |
| *leuk* | 0 | 1 | 12 | 0 | 0 | 6 | 4 | 3 | 276 | 0 | 0 | 1 | 0 | 1 |
| *kid* | 5 | 3 | 10 | 1 | 0 | 4 | 6 | 2 | 0 | 219 | 0 | 1 | 0 | 0 |
| *pan* | 0 | 1 | 1 | 0 | 0 | 0 | 2 | 0 | 0 | 0 | 34 | 0 | 0 | 0 |
| *oral* | 0 | 1 | 8 | 0 | 2 | 3 | 3 | 1 | 1 | 1 | 0 | 184 | 1 | 1 |
| *nhl* | 1 | 1 | 12 | 2 | 0 | 5 | 6 | 1 | 0 | 0 | 0 | 1 | 383 | 0 |
| *lung* | 2 | 7 | 26 | 8 | 1 | 5 | 3 | 4 | 1 | 0 | 0 | 1 | 0 | 174 |

**Supplementary Table 3.** AUCs of ROC curves based on a logistic regression model with specific cancer diagnosis as a dependent variable and age as well as other cancer diagnoses as independent variables. No cancer PRS were included in these models. Other cancers are chosen after both-way stepwise logistic regression keeping age fixed. We call this model the final model.

| Cancer | Cases | Controls | AUC (age) | AUC (final) | Cancers in the Final Model |
| --- | --- | --- | --- | --- | --- |
| **MALES** |  |  |  |  |  |
| Bladder | 835 | 196731 | 0.7020 | 0.7123 | prostate, kidney, testicular, lung |
| Colon | 835 | 196731 | 0.6991 | 0.7074 | lung, pancreas, leukemia |
| Thyroid | 90 | 197476 | 0.5090 | 0.5090 | - |
| Melanoma | 1492 | 196074 | 0.6015 | 0.6037 | testicular, NHL |
| Kidney | 377 | 197189 | 0.6317 | 0.6565 | lung, bladder, prostate, testicular, pancreas |
| Leukemia | 397 | 197169 | 0.6161 | 0.6217 | NHL, colon |
| Lung | 227 | 197339 | 0.6758 | 0.7409 | colon, kidney, testicular, oral, bladder, prostate |
| NHL | 513 | 197053 | 0.5836 | 0.5891 | leukemia, testicular, melanoma |
| Oral | 293 | 197273 | 0.5959 | 0.5969 | lung |
| Pancreas | 49 | 197517 | 0.6413 | 0.6705 | colon, prostate, kidney |
| Prostate | 3700 | 193866 | 0.7476 | 0.7494 | bladder, kidney, pancreas |
| Testicular | 774 | 196792 | 0.5764 | 0.5872 | lung, bladder, NHL, melanoma, kidney |
| **FEMALES** |  |  |  |  |  |
| Bladder | 302 | 232761 | 0.6962 | 0.7068 | kidney, lung, colon, cervix, melanoma |
| Colon | 638 | 232425 | 0.6929 | 0.7041 | lung, ovarian, endometrium, kidney, bladder, pancreas, breast, cervix |
| Thyroid | 330 | 232733 | 0.5212 | 0.5248 | oral, ovarian |
| Melanoma | 2101 | 230962 | 0.5689 | 0.5702 | kidney, pancreas, bladder |
| Kidney | 219 | 232844 | 0.6299 | 0.6430 | bladder, melanoma, colon |
| Leukemia | 276 | 232787 | 0.5921 | 0.5921 | - |
| Lung | 174 | 232889 | 0.6631 | 0.7229 | ovarian, breast, colon, bladder, endometrium, oral |
| NHL | 383 | 232680 | 0.5814 | 0.5859 | breast |
| Oral | 184 | 232879 | 0.5879 | 0.5907 | thyroid |
| Pancreas | 34 | 233029 | 0.5390 | 0.5824 | melanoma, colon |
| Cervix | 3569 | 229494 | 0.5448 | 0.5521 | endometrium, breast, ovarian, bladder, colon |
| Ovarian | 755 | 232308 | 0.5983 | 0.6204 | endometrium, lung, colon, breast, cervix, thyroid, leukemia |
| Breast | 10091 | 222972 | 0.6226 | 0.6244 | lung, ovarian, cervix, endometrium, NHL, colon |
| Endometrium | 1137 | 231926 | 0.6671 | 0.6821 | ovarian, cervix, colon, breast, lung |

**Supplementary Table 4.**
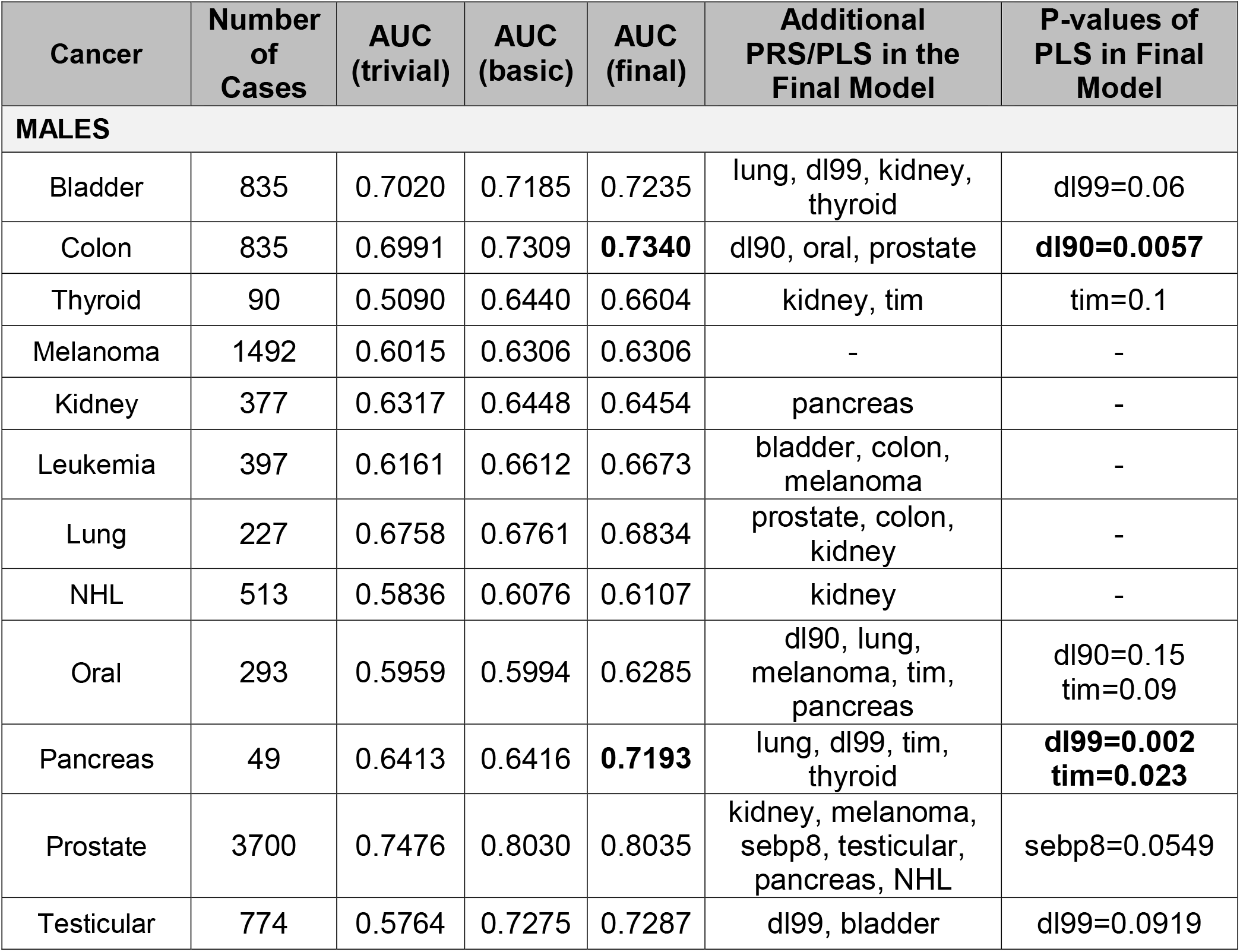
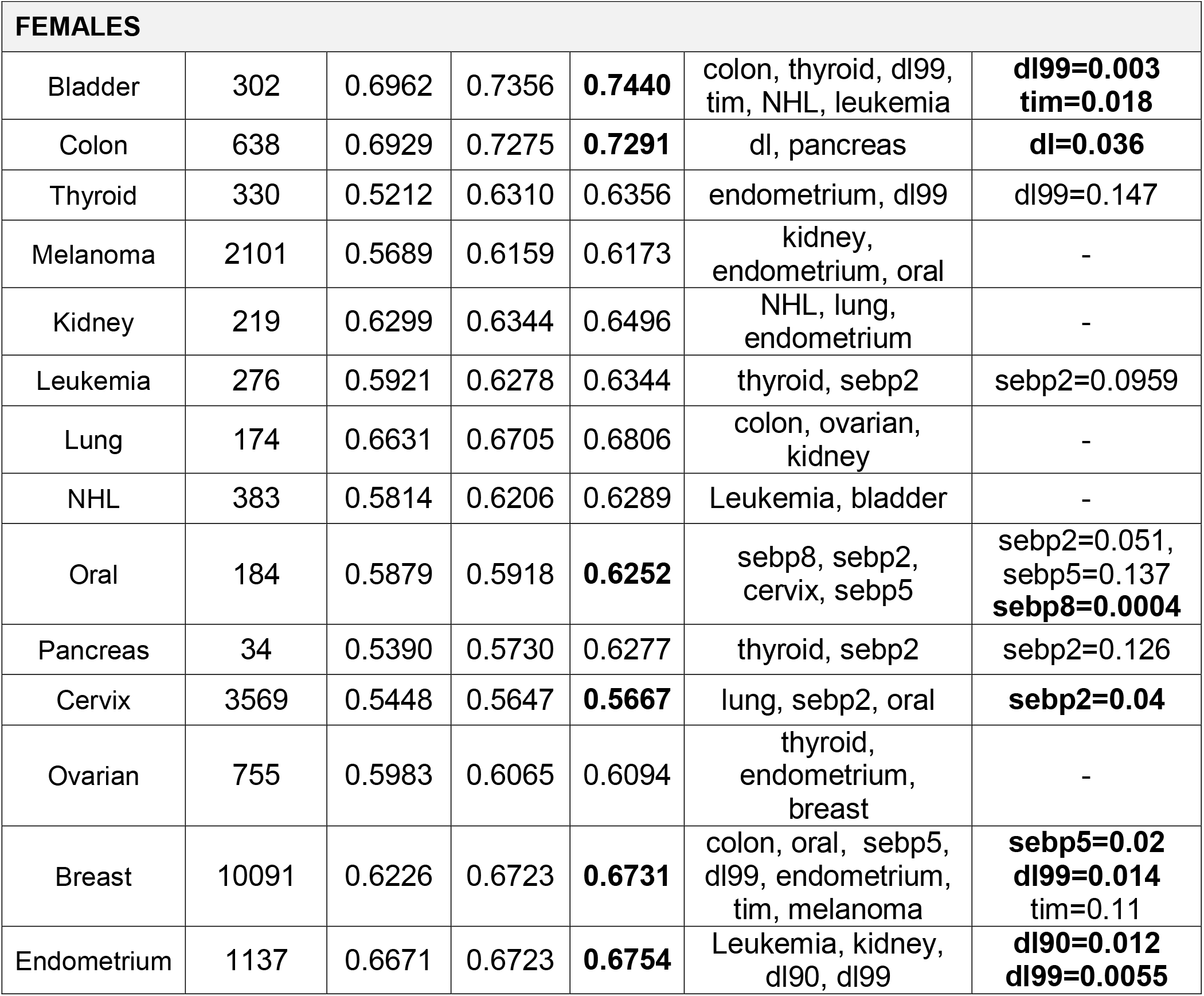
AUC values derived for ROC analyses of the results of logistic regression modeling with specific cancer diagnosis as a dependent variable and the corresponding PRS for that cancer, PRS of the other cancers, age, and the different PLSs as independent variables. We fit three models to determine the significance of the contribution of PLS: a ‘trivial model’ with just age as an independent variable; a ‘basic’ model with age and the cancer-specific PRS as independent variables; and a ‘final’ model in which age, the cancer-specific PRS, the PRSs of the other cancers, and PLSs were independent variables subjected to forward and backward stepwise logistic regression with age fixed in all models. The bolded entries included a significant PLS effect.

**Supplementary Table 5.** Correlations of Cancer-PRS vs Longevity Scores. The table shows p-values for significant correlations (p-value < 0.05). Bold type-face indicates a statistically significant negative correlation or positive correlation between the specific longevity score and cancer type.

| Cancer | dl<br>(p-val) | dl90<br>(p-val) | dl99<br>(p-val) | tim<br>(p-val) | seb10<br>(p-val) | sebp2<br>(p-val) | sebp5<br>(p-val) | sebp8<br>(p-val) |
| --- | --- | --- | --- | --- | --- | --- | --- | --- |
| Bladder | 0.0023 | 0.0015 | 0.0024 | -0.0004 | 0.0011 | 0.0017 | -0.0017 | 0.0002 |
| Colon | 0.0009 | **0.0047**<br>**(0.002)** | 0.0008 | **0.0349**<br>**($<10^{-100}$)** | -0.0004 | 0.0002 | **0.0300**<br>**($<10^{-50}$)** | -0.0003 |
| Thyroid | **-0.0033**<br>**(0.028)** | -0.002 | -0.0033<br>(0.032) | -0.0029 | -0.0025 | 0.0015 | -0.0003 | -0.0021 |
| Melanoma | **-0.0032**<br>**(0.033)** | -0.0014 | -0.0027 | -0.0011 | **-0.0041**<br>**(0.0068)** | **-0.0067**<br>**($10^{-5}$)** | -0.0013 | -0.0026 |
| Kidney | -0.0010 | -0.0023 | -0.0014 | -0.0007 | 0.0001 | 0.0017 | -0.0018 | -0.0009 |
| Leukemia | 0.0002 | -0.0003 | -0.0001 | **-0.0061**<br>**(0.00006)** | 0.0005 | **-0.0074**<br>**($<10^{-5}$)** | **-0.0042**<br>**(0.0055)** | 0.0015 |
| Lung | 0.0004 | -0.0001 | 0.0005 | **-0.1925**<br>**($<10^{-100}$)** | **-0.0134**<br>**($<10^{-17}$)** | **-0.0081**<br>**($<10^{-6}$)** | **0.0166**<br>**($<10^{-27}$)** | 0.0007 |
| NHL | 0.0002 | 0.0002 | 0.0001 | **-0.0243**<br>**($<10^{-50}$)** | -0.0005 | -0.0004 | 0.0006 | -0.0005 |
| Oral | -0.0002 | 0.0005 | -0.0002 | **0.0054**<br>**(0.0004)** | **-0.0041**<br>**(0.0077)** | **0.0103**<br>**($<10^{-10}$)** | **0.0551**<br>**($<10^{-100}$)** | 0.0001 |
| Pancreas | 0.0005 | 0.0012 | 0.0003 | **-0.0311**<br>**($<10^{-50}$)** | 0.0008 | -0.0009 | 0.0012 | 0.0003 |
| Prostate | 0.0008 | -0.0007 | 0.0008 | **-0.0124**<br>**($<10^{-15}$)** | -0.0021 | **-0.0054**<br>**(0.00035)** | -0.0001 | -0.0006 |
| Testicular | -0.0004 | -0.0006 | -0.0004 | 0.0018 | 0.0002 | **0.0306**<br>**($<10^{-50}$)** | 0.0030 | 0.0004 |
| Cervix | **-0.0043**<br>**(0.005)** | **-0.0035**<br>**(0.02)** | **-0.0039**<br>**(0.01)** | **-0.0535**<br>**($<10^{-100}$)** | -0.0028 | **-0.0057**<br>**(0.0002)** | -0.0001 | -0.0023 |
| Ovarian | -0.0013 | -0.0025 | -0.0013 | **-0.0035**<br>**(0.023)** | -0.0014 | -0.0016 | -0.0012 | -0.0014 |
| Breast | 0.0002 | 0.0001 | 0.0003 | -0.0029 | -0.0006 | 0.0001 | -0.0002 | -0.0005 |
| Endometrium | 0.0009 | 0.0008 | 0.0012 | -0.0005 | 0.0015 | **0.0110**<br>**($<10^{-12}$)** | 0.0023 | 0.0002 |

**Supplementary Table 6.** Matrix with number of SNPs in the summary statistics used to generate cancer PRSs (in the diagonal) and number of intersecting SNPs across the cancers (in corresponding non-diagonal positions).

|  | bladder | breast | cervix | colon | endom | kid | leuk | lung | mela | nhl | oral | ova | pan | prost | testi | thy |
| --- | --- | --- | --- | --- | --- | --- | --- | --- | --- | --- | --- | --- | --- | --- | --- | --- |
| *bladder* | 15 | 0 | 0 | 1 | 0 | 0 | 1 | 0 | 1 | 0 | 0 | 0 | 1 | 0 | 0 | 0 |
| *breast* | 0 | 177 | 0 | 0 | 0 | 1 | 0 | 0 | 0 | 0 | 0 | 0 | 0 | 2 | 0 | 0 |
| *cervix* | 0 | 0 | 10 | 0 | 0 | 0 | 0 | 0 | 0 | 0 | 0 | 0 | 0 | 0 | 0 | 0 |
| *colon* | 1 | 0 | 0 | 103 | 0 | 0 | 1 | 0 | 0 | 0 | 0 | 0 | 0 | 0 | 0 | 0 |
| *endom* | 0 | 0 | 0 | 0 | 9 | 0 | 0 | 0 | 0 | 0 | 0 | 0 | 0 | 0 | 0 | 0 |
| *kid* | 0 | 1 | 0 | 0 | 0 | 19 | 0 | 0 | 0 | 0 | 0 | 0 | 0 | 0 | 0 | 0 |
| *leuk* | 1 | 0 | 0 | 1 | 0 | 0 | 75 | 1 | 0 | 0 | 0 | 2 | 0 | 0 | 0 | 0 |
| *lung* | 0 | 0 | 0 | 0 | 0 | 0 | 1 | 71 | 0 | 0 | 0 | 1 | 0 | 0 | 1 | 0 |
| *mela* | 1 | 0 | 0 | 0 | 0 | 0 | 0 | 0 | 23 | 0 | 0 | 0 | 1 | 1 | 0 | 0 |
| *nhl* | 0 | 0 | 0 | 0 | 0 | 0 | 0 | 0 | 0 | 19 | 0 | 0 | 0 | 0 | 0 | 0 |
| *oral* | 0 | 0 | 0 | 0 | 0 | 0 | 0 | 0 | 0 | 0 | 13 | 0 | 0 | 0 | 0 | 0 |
| *ova* | 0 | 0 | 0 | 0 | 0 | 0 | 2 | 1 | 0 | 0 | 0 | 31 | 0 | 0 | 0 | 1 |
| *pan* | 1 | 0 | 0 | 0 | 0 | 0 | 0 | 0 | 1 | 0 | 0 | 0 | 21 | 0 | 0 | 0 |
| *prost* | 0 | 2 | 0 | 0 | 0 | 0 | 0 | 0 | 1 | 0 | 0 | 0 | 0 | 158 | 0 | 0 |
| *testi* | 0 | 0 | 0 | 0 | 0 | 0 | 0 | 1 | 0 | 0 | 0 | 0 | 0 | 0 | 51 | 0 |
| *thy* | 0 | 0 | 0 | 0 | 0 | 0 | 0 | 0 | 0 | 0 | 0 | 1 | 0 | 0 | 0 | 10 |

**Supplementary Table 7.**
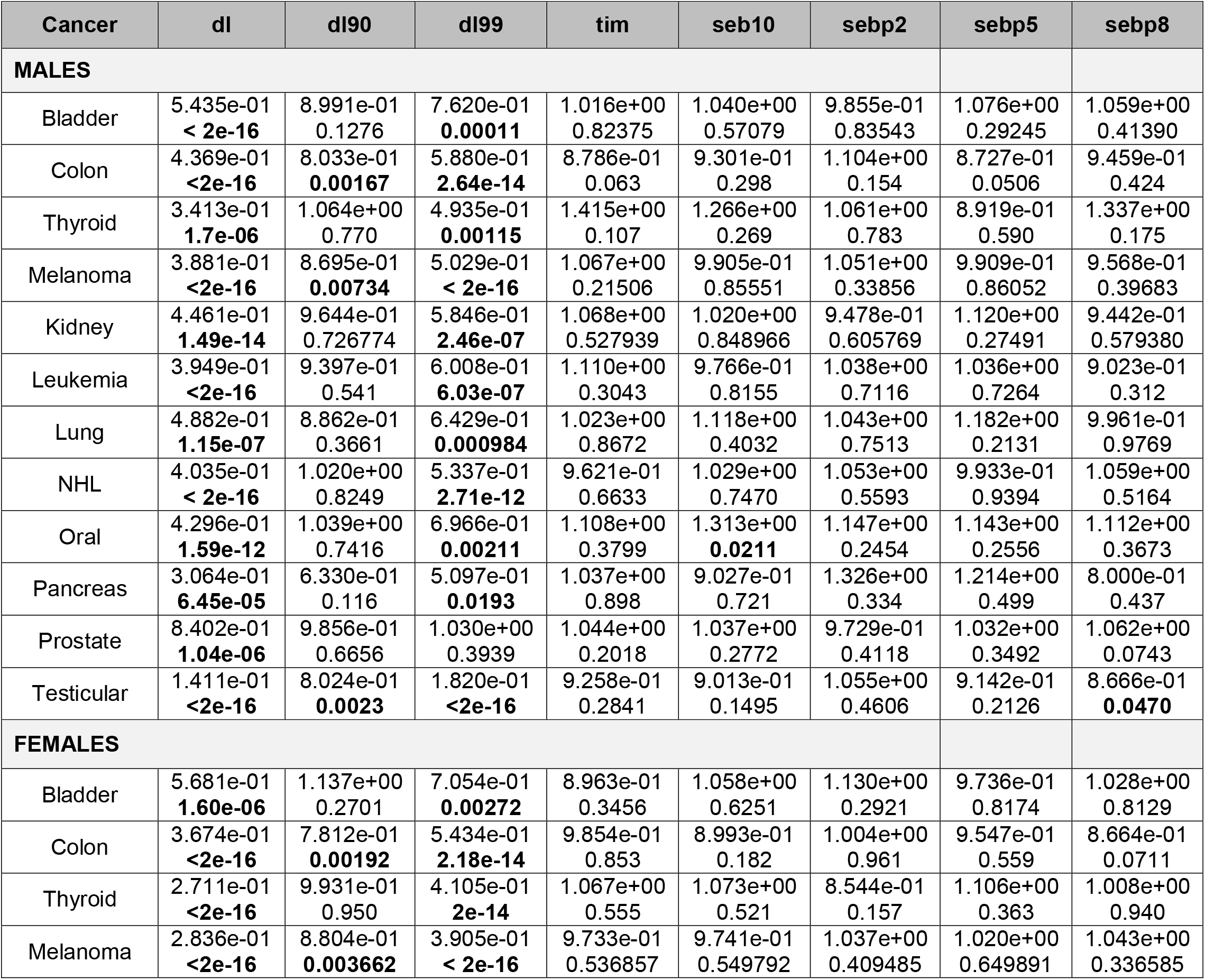
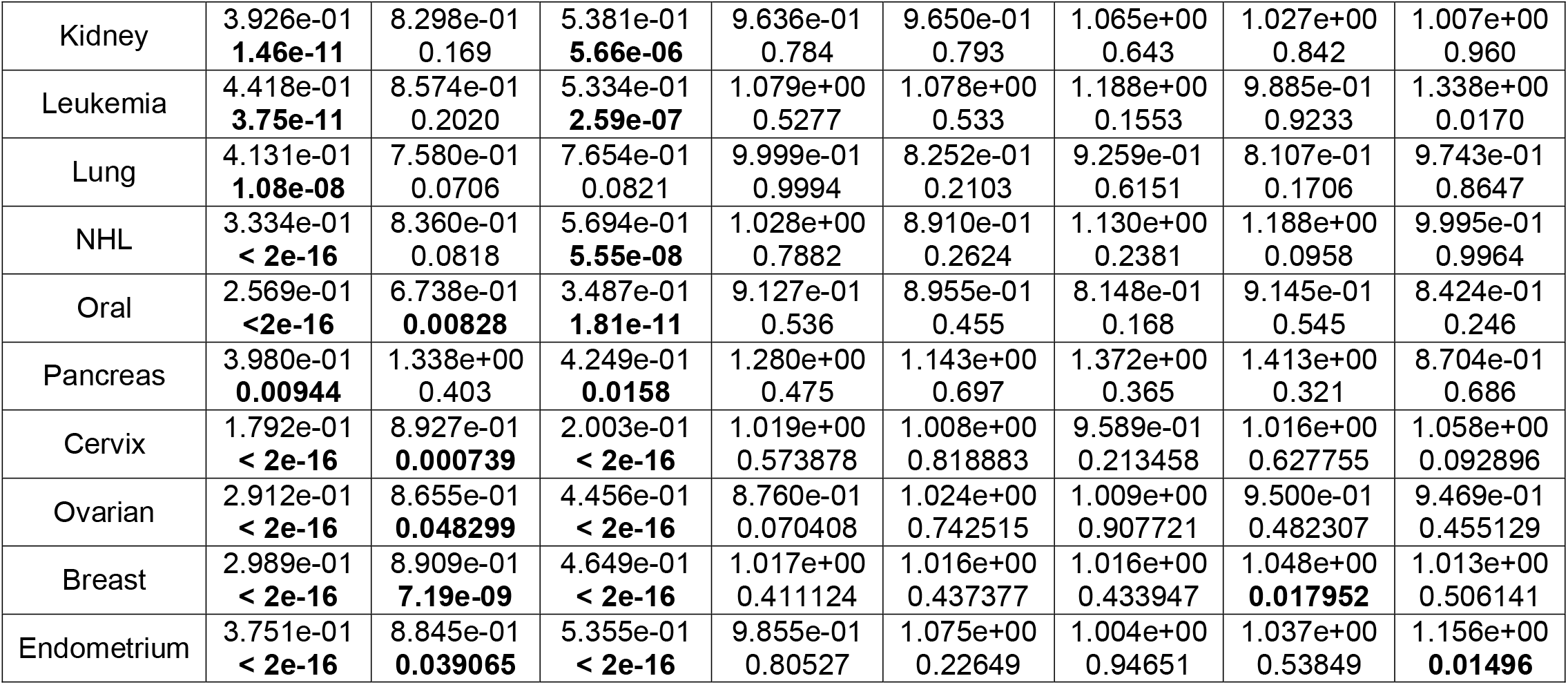
Cox proportional hazards models testing the difference in the upper and lower 50^th^ percentiles of the 8 different PLS against specific cancer age-of-onsets. The first 10 PCs of the genome-wide GRM for individuals considered in the study as well as the cancer PRS for the specific cancer whose onset is of focus were included as covariates. The upper entries provide the estimated hazard ratios for those in the upper 50^th^ percentile of the PLS distribution and the lower entries provide the p-value for the difference from those in the lower 50^th^ percentiles. Bold entries have p<0.05.

| Cancer | dl | dl90 | dl99 | tim | seb10 | sebp2 | sebp5 | sebp8 |
| --- | --- | --- | --- | --- | --- | --- | --- | --- |
| **MALES** |  |  |  |  |  |  |  |  |
| Bladder | 5.435e-01<br>**< 2e-16** | 8.991e-01<br>0.1276 | 7.620e-01<br>**0.00011** | 1.016e+00<br>0.82375 | 1.040e+00<br>0.57079 | 9.855e-01<br>0.83543 | 1.076e+00<br>0.29245 | 1.059e+00<br>0.41390 |
| Colon | 4.369e-01<br>**<2e-16** | 8.033e-01<br>**0.00167** | 5.880e-01<br>**2.64e-14** | 8.786e-01<br>0.063 | 9.301e-01<br>0.298 | 1.104e+00<br>0.154 | 8.727e-01<br>0.0506 | 9.459e-01<br>0.424 |
| Thyroid | 3.413e-01<br>**1.7e-06** | 1.064e+00<br>0.770 | 4.935e-01<br>**0.00115** | 1.415e+00<br>0.107 | 1.266e+00<br>0.269 | 1.061e+00<br>0.783 | 8.919e-01<br>0.590 | 1.337e+00<br>0.175 |
| Melanoma | 3.881e-01<br>**<2e-16** | 8.695e-01<br>**0.00734** | 5.029e-01<br>**< 2e-16** | 1.067e+00<br>0.21506 | 9.905e-01<br>0.85551 | 1.051e+00<br>0.33856 | 9.909e-01<br>0.86052 | 9.568e-01<br>0.39683 |
| Kidney | 4.461e-01<br>**1.49e-14** | 9.644e-01<br>0.726774 | 5.846e-01<br>**2.46e-07** | 1.068e+00<br>0.527939 | 1.020e+00<br>0.848966 | 9.478e-01<br>0.605769 | 1.120e+00<br>0.27491 | 9.442e-01<br>0.579380 |
| Leukemia | 3.949e-01<br>**<2e-16** | 9.397e-01<br>0.541 | 6.008e-01<br>**6.03e-07** | 1.110e+00<br>0.3043 | 9.766e-01<br>0.8155 | 1.038e+00<br>0.7116 | 1.036e+00<br>0.7264 | 9.023e-01<br>0.312 |
| Lung | 4.882e-01<br>**1.15e-07** | 8.862e-01<br>0.3661 | 6.429e-01<br>**0.000984** | 1.023e+00<br>0.8672 | 1.118e+00<br>0.4032 | 1.043e+00<br>0.7513 | 1.182e+00<br>0.2131 | 9.961e-01<br>0.9769 |
| NHL | 4.035e-01<br>**< 2e-16** | 1.020e+00<br>0.8249 | 5.337e-01<br>**2.71e-12** | 9.621e-01<br>0.6633 | 1.029e+00<br>0.7470 | 1.053e+00<br>0.5593 | 9.933e-01<br>0.9394 | 1.059e+00<br>0.5164 |
| Oral | 4.296e-01<br>**1.59e-12** | 1.039e+00<br>0.7416 | 6.966e-01<br>**0.00211** | 1.108e+00<br>0.3799 | 1.313e+00<br>**0.0211** | 1.147e+00<br>0.2454 | 1.143e+00<br>0.2556 | 1.112e+00<br>0.3673 |
| Pancreas | 3.064e-01<br>**6.45e-05** | 6.330e-01<br>0.116 | 5.097e-01<br>**0.0193** | 1.037e+00<br>0.898 | 9.027e-01<br>0.721 | 1.326e+00<br>0.334 | 1.214e+00<br>0.499 | 8.000e-01<br>0.437 |
| Prostate | 8.402e-01<br>**1.04e-06** | 9.856e-01<br>0.6656 | 1.030e+00<br>0.3939 | 1.044e+00<br>0.2018 | 1.037e+00<br>0.2772 | 9.729e-01<br>0.4118 | 1.032e+00<br>0.3492 | 1.062e+00<br>0.0743 |
| Testicular | 1.411e-01<br>**<2e-16** | 8.024e-01<br>**0.0023** | 1.820e-01<br>**<2e-16** | 9.258e-01<br>0.2841 | 9.013e-01<br>0.1495 | 1.055e+00<br>0.4606 | 9.142e-01<br>0.2126 | 8.666e-01<br>**0.0470** |
| **FEMALES** |  |  |  |  |  |  |  |  |
| Bladder | 5.681e-01<br>**1.60e-06** | 1.137e+00<br>0.2701 | 7.054e-01<br>**0.00272** | 8.963e-01<br>0.3456 | 1.058e+00<br>0.6251 | 1.130e+00<br>0.2921 | 9.736e-01<br>0.8174 | 1.028e+00<br>0.8129 |
| Colon | 3.674e-01<br>**<2e-16** | 7.812e-01<br>**0.00192** | 5.434e-01<br>**2.18e-14** | 9.854e-01<br>0.853 | 8.993e-01<br>0.182 | 1.004e+00<br>0.961 | 9.547e-01<br>0.559 | 8.664e-01<br>0.0711 |
| Thyroid | 2.711e-01<br>**<2e-16** | 9.931e-01<br>0.950 | 4.105e-01<br>**2e-14** | 1.067e+00<br>0.555 | 1.073e+00<br>0.521 | 8.544e-01<br>0.157 | 1.106e+00<br>0.363 | 1.008e+00<br>0.940 |
| Melanoma | 2.836e-01<br>**<2e-16** | 8.804e-01<br>**0.003662** | 3.905e-01<br>**< 2e-16** | 9.733e-01<br>0.536857 | 9.741e-01<br>0.549792 | 1.037e+00<br>0.409485 | 1.020e+00<br>0.649891 | 1.043e+00<br>0.336585 |
| Kidney | 3.926e-01<br>**1.46e-11** | 8.298e-01<br>0.169 | 5.381e-01<br>**5.66e-06** | 9.636e-01<br>0.784 | 9.650e-01<br>0.793 | 1.065e+00<br>0.643 | 1.027e+00<br>0.842 | 1.007e+00<br>0.960 |
| Leukemia | 4.418e-01<br>**3.75e-11** | 8.574e-01<br>0.2020 | 5.334e-01<br>**2.59e-07** | 1.079e+00<br>0.5277 | 1.078e+00<br>0.533 | 1.188e+00<br>0.1553 | 9.885e-01<br>0.9233 | 1.338e+00<br>0.0170 |
| Lung | 4.131e-01<br>**1.08e-08** | 7.580e-01<br>0.0706 | 7.654e-01<br>0.0821 | 9.999e-01<br>0.9994 | 8.252e-01<br>0.2103 | 9.259e-01<br>0.6151 | 8.107e-01<br>0.1706 | 9.743e-01<br>0.8647 |
| NHL | 3.334e-01<br>**< 2e-16** | 8.360e-01<br>0.0818 | 5.694e-01<br>**5.55e-08** | 1.028e+00<br>0.7882 | 8.910e-01<br>0.2624 | 1.130e+00<br>0.2381 | 1.188e+00<br>0.0958 | 9.995e-01<br>0.9964 |
| Oral | 2.569e-01<br>**<2e-16** | 6.738e-01<br>**0.00828** | 3.487e-01<br>**1.81e-11** | 9.127e-01<br>0.536 | 8.955e-01<br>0.455 | 8.148e-01<br>0.168 | 9.145e-01<br>0.545 | 8.424e-01<br>0.246 |
| Pancreas | 3.980e-01<br>**0.00944** | 1.338e+00<br>0.403 | 4.249e-01<br>**0.0158** | 1.280e+00<br>0.475 | 1.143e+00<br>0.697 | 1.372e+00<br>0.365 | 1.413e+00<br>0.321 | 8.704e-01<br>0.686 |
| Cervix | 1.792e-01<br>**< 2e-16** | 8.927e-01<br>**0.000739** | 2.003e-01<br>**< 2e-16** | 1.019e+00<br>0.573878 | 1.008e+00<br>0.818883 | 9.589e-01<br>0.213458 | 1.016e+00<br>0.627755 | 1.058e+00<br>0.092896 |
| Ovarian | 2.912e-01<br>**< 2e-16** | 8.655e-01<br>**0.048299** | 4.456e-01<br>**< 2e-16** | 8.760e-01<br>0.070408 | 1.024e+00<br>0.742515 | 1.009e+00<br>0.907721 | 9.500e-01<br>0.482307 | 9.469e-01<br>0.455129 |
| Breast | 2.989e-01<br>**< 2e-16** | 8.909e-01<br>**7.19e-09** | 4.649e-01<br>**< 2e-16** | 1.017e+00<br>0.411124 | 1.016e+00<br>0.437377 | 1.016e+00<br>0.433947 | 1.048e+00<br>**0.017952** | 1.013e+00<br>0.506141 |
| Endometrium | 3.751e-01<br>**< 2e-16** | 8.845e-01<br>**0.039065** | 5.355e-01<br>**< 2e-16** | 9.855e-01<br>0.80527 | 1.075e+00<br>0.22649 | 1.004e+00<br>0.94651 | 1.037e+00<br>0.53849 | 1.156e+00<br>**0.01496** |

**Supplementary Table 8.** Kaplan-Meier analyses for each cancer and each PLS. Values represent the rounded p-values from comparison of the two survival curves based on the upper and lower halves (50^th^ percentiles) of PLS. The values are tabulated for each of the PLS and for each of the 16 cancers as well as ‘any cancer’. PLSs ‘dl’, ‘dl90’ and ‘dl99’ show consistently significant differences between the upper and lower 50^th^ percentile curves.

| Cancer | tim | dl | dl90 | dl99 | seb10 | sebp2 | sebp5 | sebp8 |
| --- | --- | --- | --- | --- | --- | --- | --- | --- |
| **MALES** |  |  |  |  |  |  |  |  |
| Any | 0.1 | <2e-16 | 7e-05 | <2e-16 | 0.4 | 0.6 | 0.06 | 0.8 |
| Bladder | 0.9 | <2e-16 | 0.2 | 4e-04 | 0.8 | 0.7 | 0.3 | 0.4 |
| Colon | 0.1 | <2e-16 | 0.003 | 1e-13 | 0.2 | 0.2 | 0.1 | 0.5 |
| Thyroid | 0.1 | 7e-07 | 0.8 | 0.001 | 0.3 | 0.9 | 0.6 | 0.2 |
| Melanoma | 0.2 | <2e-16 | 0.01 | <2e-16 | 0.6 | 0.5 | 0.9 | 0.5 |
| Kidney | 0.6 | 4e-15 | 0.8 | 5e-07 | 1 | 0.6 | 0.3 | 0.6 |
| Leukemia | 0.3 | <2e-16 | 0.6 | 1e-06 | 0.7 | 0.8 | 0.7 | 0.3 |
| Lung | 1 | 6e-08 | 0.4 | 0.002 | 0.5 | 0.8 | 0.2 | 0.9 |
| NHL | 0.5 | <2e-16 | 0.8 | 9e-12 | 0.9 | 0.6 | 1 | 0.5 |
| Oral | 0.4 | 4e-13 | 0.7 | 0.004 | 0.03 | 0.3 | 0.2 | 0.3 |
| Pancreas | 0.9 | 2e-05 | 0.1 | 0.02 | 0.7 | 0.5 | 0.5 | 0.5 |
| Prostate | 0.5 | 1e-07 | 0.7 | 0.3 | 0.6 | 0.1 | 0.3 | 0.2 |
| Testicular | 0.2 | <2e-16 | 0.004 | <2e-16 | 0.07 | 0.3 | 0.2 | 0.05 |
| **FEMALES** |  |  |  |  |  |  |  |  |
| Any | 0.9 | <2e-16 | <2e-16 | <2e-16 | 0.1 | 0.2 | 0.02 | 0.04 |
| Bladder | 0.3 | 2e-06 | 0.2 | 0.005 | 0.8 | 0.4 | 0.8 | 0.8 |
| Colon | 0.9 | <2e-16 | 0.003 | 8e-14 | 0.1 | 0.8 | 0.8 | 0.09 |
| Thyroid | 0.6 | <2e-16 | 1 | 2e-14 | 0.7 | 0.1 | 0.3 | 0.9 |
| Melanoma | 0.5 | <2e-16 | 0.006 | <2e-16 | 0.2 | 0.9 | 0.6 | 0.3 |
| Kidney | 0.8 | 5e-12 | 0.2 | 7e-06 | 0.7 | 0.7 | 0.8 | 0.9 |
| Leukemia | 0.6 | 2e-11 | 0.2 | 5e-07 | 0.6 | 0.2 | 0.9 | 0.01 |
| Lung | 0.7 | 4e-09 | 0.08 | 0.1 | 0.2 | 0.5 | 0.2 | 0.9 |
| NHL | 0.9 | <2e-16 | 0.1 | 1e-07 | 0.2 | 0.4 | 0.09 | 0.9 |
| Oral | 0.5 | <2e-16 | 0.009 | 9e-12 | 0.4 | 0.1 | 0.6 | 0.3 |
| Pancreas | 0.5 | 0.008 | 0.4 | 0.02 | 0.7 | 0.5 | 0.3 | 0.7 |
| Cervix | 1 | <2e-16 | 0.002 | <2e-16 | 0.6 | 0.02 | 0.5 | 0.05 |
| Ovarian | 0.06 | <2e-16 | 0.06 | <2e-16 | 1 | 0.8 | 0.5 | 0.5 |
| Breast | 0.6 | <2e-16 | 5e-08 | <2e-16 | 0.8 | 0.6 | 0.006 | 0.2 |
| Endometrium | 0.8 | <2e-16 | 0.06 | <2e-16 | 0.4 | 0.8 | 0.5 | 0.01 |

**Supplementary Table 9.**
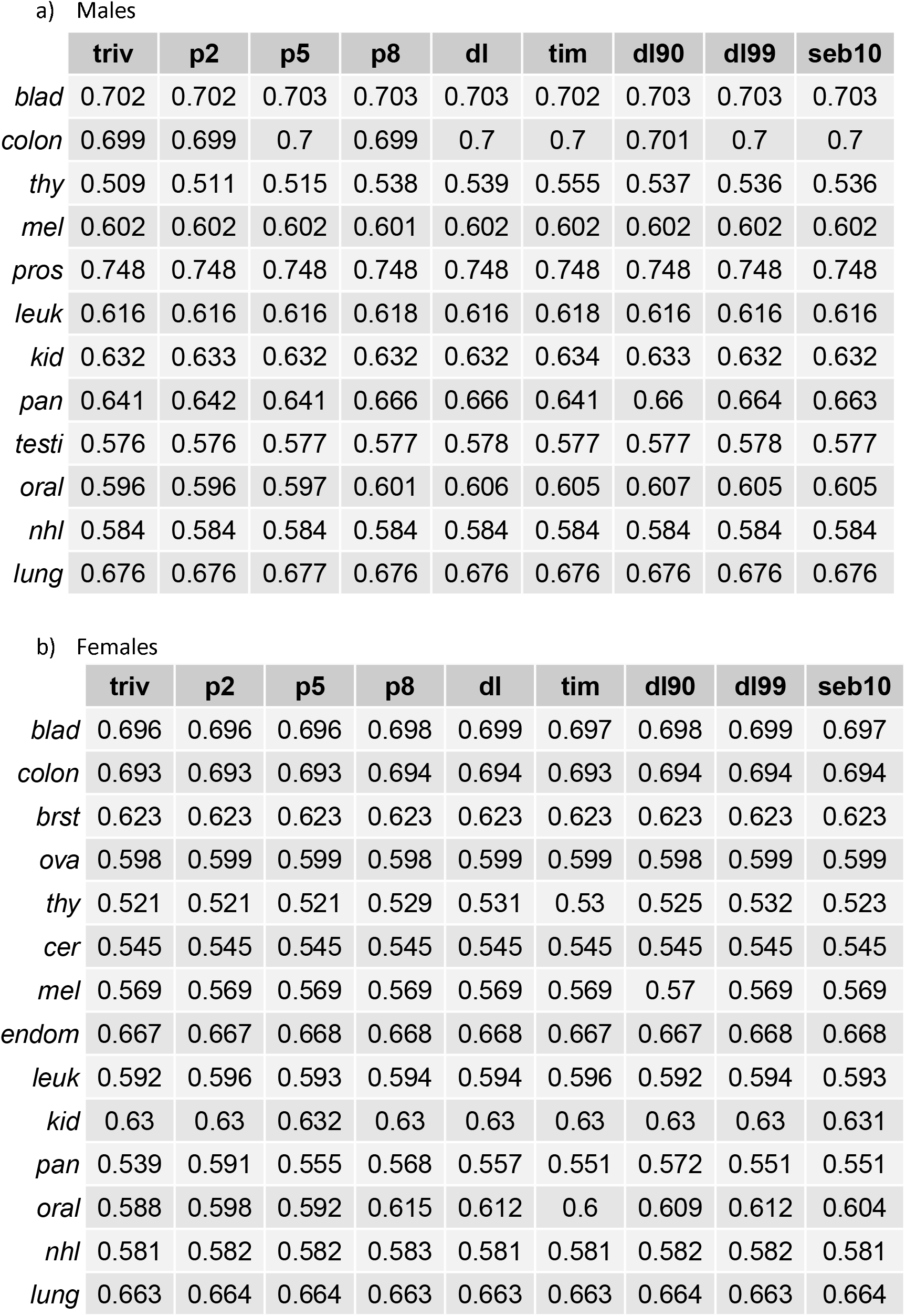
AUCs of ROC of each cancer prediction with age (triv) and AUCs with age plus each of the PLSs.

a) Males
|  | **triv** | **p2** | **p5** | **p8** | **dl** | **tim** | **dl90** | **dl99** | **seb10** |
| --- | --- | --- | --- | --- | --- | --- | --- | --- | --- |
| *blad* | 0.702 | 0.702 | 0.703 | 0.703 | 0.703 | 0.702 | 0.703 | 0.703 | 0.703 |
| *colon* | 0.699 | 0.699 | 0.7 | 0.699 | 0.7 | 0.7 | 0.701 | 0.7 | 0.7 |
| *thy* | 0.509 | 0.511 | 0.515 | 0.538 | 0.539 | 0.555 | 0.537 | 0.536 | 0.536 |
| *mel* | 0.602 | 0.602 | 0.602 | 0.601 | 0.602 | 0.602 | 0.602 | 0.602 | 0.602 |
| *pros* | 0.748 | 0.748 | 0.748 | 0.748 | 0.748 | 0.748 | 0.748 | 0.748 | 0.748 |
| *leuk* | 0.616 | 0.616 | 0.616 | 0.618 | 0.616 | 0.618 | 0.616 | 0.616 | 0.616 |
| *kid* | 0.632 | 0.633 | 0.632 | 0.632 | 0.632 | 0.634 | 0.633 | 0.632 | 0.632 |
| *pan* | 0.641 | 0.642 | 0.641 | 0.666 | 0.666 | 0.641 | 0.66 | 0.664 | 0.663 |
| *testi* | 0.576 | 0.576 | 0.577 | 0.577 | 0.578 | 0.577 | 0.577 | 0.578 | 0.577 |
| *oral* | 0.596 | 0.596 | 0.597 | 0.601 | 0.606 | 0.605 | 0.607 | 0.605 | 0.605 |
| *nhl* | 0.584 | 0.584 | 0.584 | 0.584 | 0.584 | 0.584 | 0.584 | 0.584 | 0.584 |
| *lung* | 0.676 | 0.676 | 0.677 | 0.676 | 0.676 | 0.676 | 0.676 | 0.676 | 0.676 |

b) Females
|  | **triv** | **p2** | **p5** | **p8** | **dl** | **tim** | **dl90** | **dl99** | **seb10** |
| --- | --- | --- | --- | --- | --- | --- | --- | --- | --- |
| *blad* | 0.696 | 0.696 | 0.696 | 0.698 | 0.699 | 0.697 | 0.698 | 0.699 | 0.697 |
| *colon* | 0.693 | 0.693 | 0.693 | 0.694 | 0.694 | 0.693 | 0.694 | 0.694 | 0.694 |
| *brst* | 0.623 | 0.623 | 0.623 | 0.623 | 0.623 | 0.623 | 0.623 | 0.623 | 0.623 |
| *ova* | 0.598 | 0.599 | 0.599 | 0.598 | 0.599 | 0.599 | 0.598 | 0.599 | 0.599 |
| *thy* | 0.521 | 0.521 | 0.521 | 0.529 | 0.531 | 0.53 | 0.525 | 0.532 | 0.523 |
| *cer* | 0.545 | 0.545 | 0.545 | 0.545 | 0.545 | 0.545 | 0.545 | 0.545 | 0.545 |
| *mel* | 0.569 | 0.569 | 0.569 | 0.569 | 0.569 | 0.569 | 0.57 | 0.569 | 0.569 |
| *endom* | 0.667 | 0.667 | 0.668 | 0.668 | 0.668 | 0.667 | 0.667 | 0.668 | 0.668 |
| *leuk* | 0.592 | 0.596 | 0.593 | 0.594 | 0.594 | 0.596 | 0.592 | 0.594 | 0.593 |
| *kid* | 0.63 | 0.63 | 0.632 | 0.63 | 0.63 | 0.63 | 0.63 | 0.63 | 0.631 |
| *pan* | 0.539 | 0.591 | 0.555 | 0.568 | 0.557 | 0.551 | 0.572 | 0.551 | 0.551 |
| *oral* | 0.588 | 0.598 | 0.592 | 0.615 | 0.612 | 0.6 | 0.609 | 0.612 | 0.604 |
| *nhl* | 0.581 | 0.582 | 0.582 | 0.583 | 0.581 | 0.581 | 0.582 | 0.582 | 0.581 |
| *lung* | 0.663 | 0.664 | 0.664 | 0.663 | 0.663 | 0.663 | 0.664 | 0.663 | 0.664 |

**Supplementary Figure 1.**
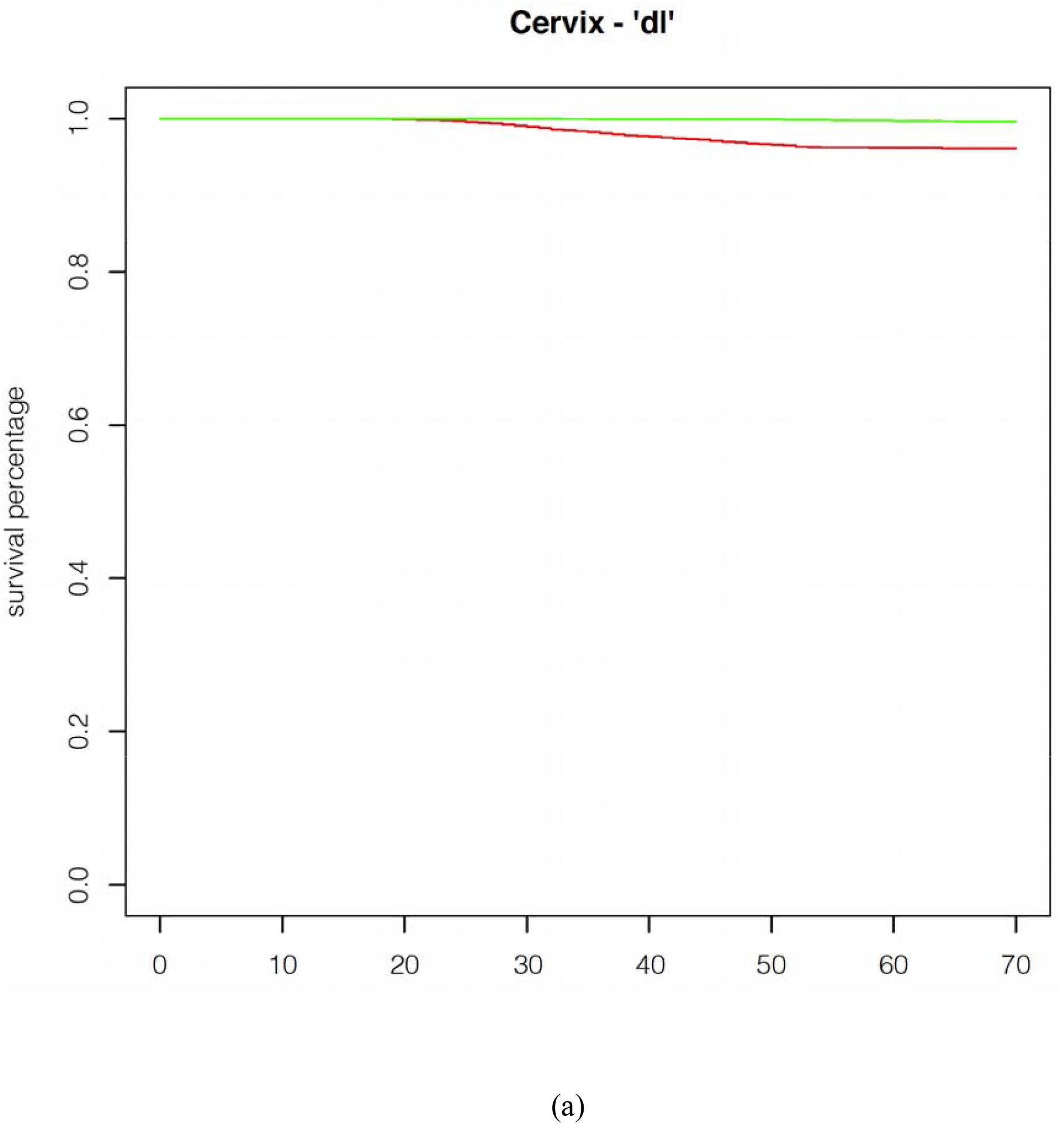
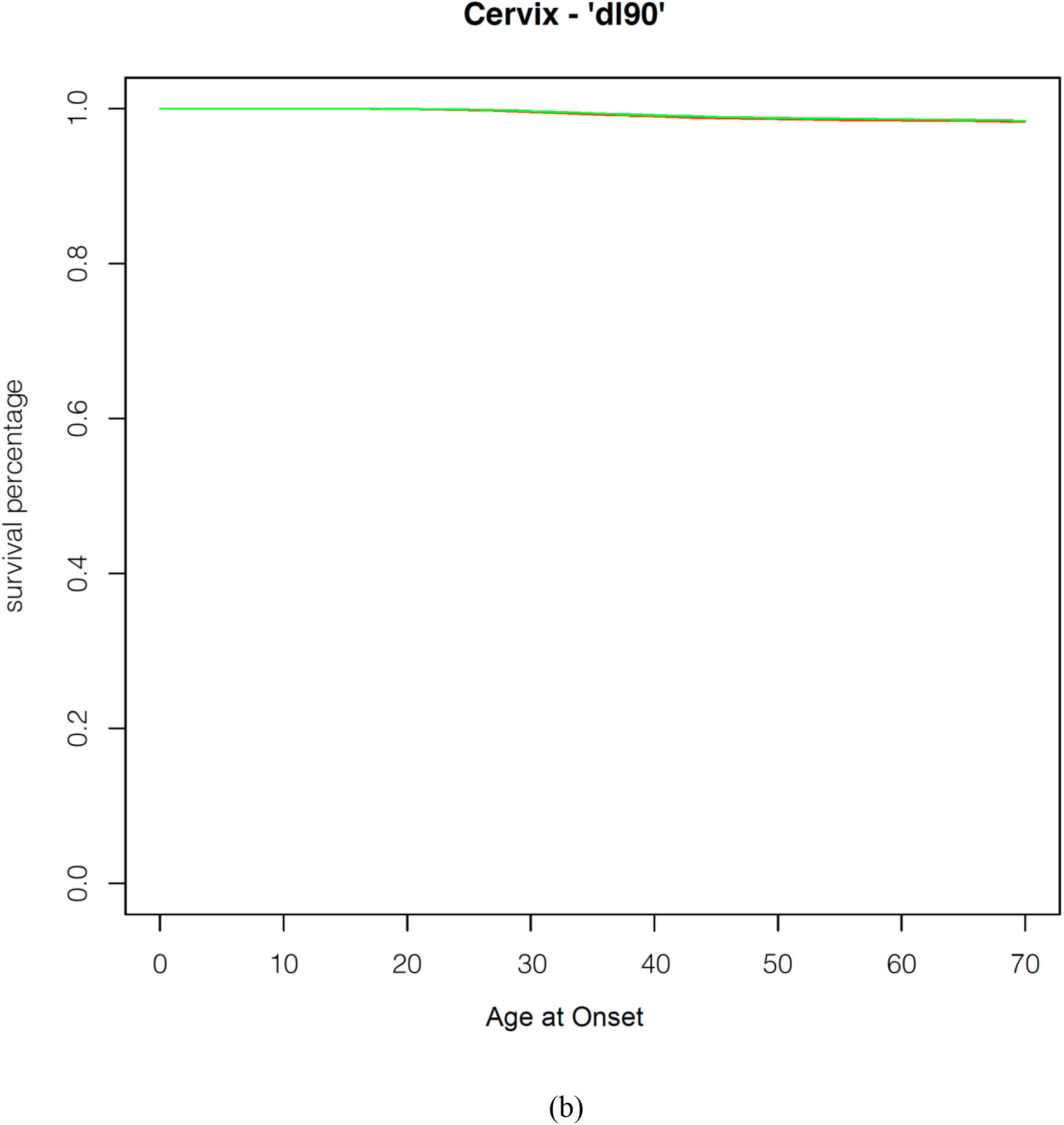
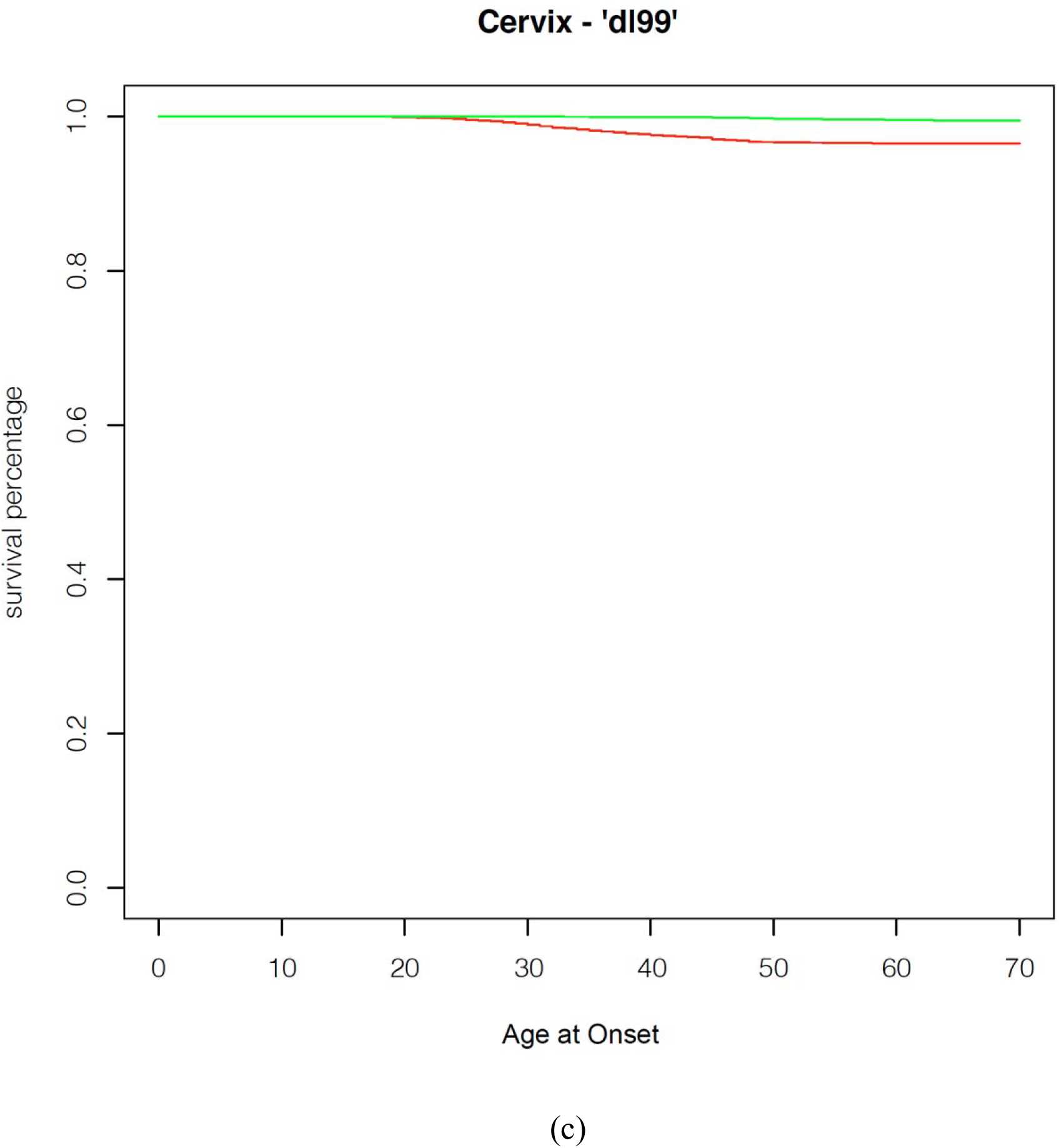
Kaplan Meier Curves showing age at onset of cervical cancer as a function of different PLSs. Green indicates individuals in the lower 10^th^ percentile of PLS and red indicates individuals in the upper 10^th^ percentile of PLS. (a) Results for PLS ‘dl’ (p<0.0001) (b) PLS ‘dl90’ (p=0.7) (c) PLS ‘dl99’ (p<0.001).

## Notes

### Competing Interest Statement

The authors have declared no competing interest.

